# Genomic epidemiology sheds light on the recent spatio-temporal dynamics of Yellow Fever virus and the spatial corridor that fueled its ongoing emergence in southern Brazil

**DOI:** 10.1101/2023.01.13.23284525

**Authors:** Marta Giovanetti, Francesco Pinotti, Camila Zanluca, Vagner Fonseca, Taishi Nakase, Andrea C. Koishi, Marcel Tscha, Guilherme Soares, Gisiane Gruber Dorl, Antônio Ernesto M.L Marques, Renato Sousa, Talita Emile Ribeiro Adelino, Joilson Xavier, Carla de Oliveira, Sandro Patroca da Silva, Natalia Rocha Guimaraes, Hegger Fritsch, Maria Angélica Mares-Guia, Flavia Levy, Pedro Henrique Passos, Vinicius Leme da Silva, Luiz Augusto Pereira, Ana Flávia Mendonça, Isabel Luana de Macêdo, Davi Emanuel Ribeiro de Sousa, Gabriela Rodrigues de Toledo Costa, Marcio Botelho de Castro, Felipe Campos de Melo Iani, Maira Alves Pereira, Karina Ribeiro Leite Jardim Cavalcante, Andre Ricardo Ribas de Freitas, Carlos Frederico Campelo de Albuquerque, Eduardo Marques Macário, Marlei Pickler Debiasi dos Anjos, Rosane Campanher Ramos, Aline Alves Scarpellini Campos, Adriano Pinter, Marcia Chame, Livia Abdalla, Irina Nastassja Riediger, Sérvio Pontes Ribeiro, Ana Isabel Bento, Tulio de Oliveira, Carla Freitas, Noely Fabiana Oliveira de Moura, Allison Fabri, Cintia Damasceno Dos Santos Rodrigues, Carolina Cardoso Dos Santos, Marco Antonio Barreto de Almeida, Edmilson dos Santos, Jader Cardoso, Douglas Adriano Augusto, Eduardo Krempser, Luís Filipe Mucci, Renata Rispoli Gatti, Sabrina Fernandes Cardoso, João Augusto Brancher Fuck, Maria Goretti David Lopes, Ivana Lucia Belmonte, Gabriela Mayoral Pedroso da Silva, Maiane Regina Ferreira Soares, Marilia de Melo Santos de Castilhos, Joseana Cardoso de Souza e Silva, Alceu Bisetto Junior, Emanuelle Gemin Pouzato, Laurina Setsuko Tanabe, Daniele Akemi Arita, Ricardo Matsuo, Josiane dos Santos Raymundo, Paula Cristina Linder Silva, Ana Santana Araújo Ferreira Silva, Sandra Samila, Glauco Carvalho, Rodrigo Stabeli, Wildo Navegantes, Luciano Andrade Moreira, Alvaro Gil A. Ferreira, Guilherme Garcia Pinheiro, Bruno Tardelli Diniz Nunes, Daniele Barbosa de Almeida Medeiros, Ana Cecília Ribeiro Cruz, Rivaldo Venâncio da Cunha, Wes Van Voorhis, Ana Maria Bispo de Filippis, Maria Almiron, Edward C. Holmes, Daniel Garkauskas Ramos, Alessandro Romano, José Lourenço, Luiz Carlos Junior Alcantara, Claudia Nunes Duarte dos Santos

## Abstract

Despite the considerable morbidity and mortality of Yellow fever virus (YFV) infections in Brazil, as well as its widespread presence in non-human primate host, our understanding of disease outbreaks is hampered by limited viral genomic data. Determining the timing and spatial corridors of YFV spread, as well as the geographic hotspots that link the endemic north of the country with epidemic extra-Amazonian regions, are central to predicting and preventing future outbreak events and epidemics. Here, we tracked the recent spread of the virus by integrating genome sequences of new YFV infections sampled from infected non-human primates and humans with both epidemiological and vector data. Through a combination of phylogenetic and epidemiological models we reconstructed the recent transmission history of YFV within different epidemic seasons in Brazil. A suitability index based on the highly domesticated *Aedes aegypti* was able to capture the seasonality of reported human infections. Spatial modelling revealed spatial hotspots with both past reporting and low vaccination coverage, which coincided with many of the largest urban centres in the Southeast. Phylodynamic analysis unravelled the circulation of three distinct YFV lineages, and provided proof of the directionality of a known spatial corridor of viral spread that connects the endemic North with the extra-Amazonian basin. This study illustrates that genomics linked with field sampling of animals and humans within a One Health framework can provide new insights into the landscape of YFV transmission, augmenting traditional approaches to infectious disease surveillance and control.

## Introduction

Yellow fever virus (YFV) is a single-stranded positive-sense RNA virus belonging to the genus *Flavivirus*, family *Flaviviridae* (Lindenbach et al., 2013). This mosquito-borne pathogen is currently endemic in tropical areas of Africa and South America. YFV can be maintained in two transmission cycles: the sylvatic (or jungle) and the urban cycles. The sylvatic cycle involves non-human primates (NHPs) and forest canopy-dwelling mosquitoes, mainly *Sabethes* sp. and *Haemagogus* sp. (Monath and Vasconcelos, 2015). While African NHPs seemingly rarely die from YFV infection, New World NHPs typically exhibit disease signs with elevated mortality rates and thus act as sentinel animals for viral circulation in the environment, in turn representing a valuable marker for epidemiological surveillance (Silva et al., 2020). Within the urban cycle, YFV is transmitted to humans by highly anthropophilic *Aedes* sp. mosquito vectors (Monath and Vasconcelos, 2015) that are typically widely distributed in urban settings in South America and Africa.

In Brazil, yellow fever transmission historically occurs within a sylvatic cycle in the Amazonian region (Vasconcelos et al., 2004). In the extra-Amazonian region, yellow fever outbreaks occur with potential infection to humans with irregular periodicity under favourable conditions for transmission, such as a build up of susceptible hosts, below threshold human vaccine coverage, high vector density, favourable temperature and rainfall, or the emergence of viral strains with potentially increased fitness advantage (Sacchetto et al., 2022; Vasconcelos et al., 1997). In recent decades, YFV re-emergence events have had a great impact on public health in Brazil (Faria et al., 2018). In 2002-2003, 2007-2009 and the during the ongoing outbreak (2016-2019 and 2020-present), there has been a spatial expansion of YFV circulation, with the virus spreading from the east towards the south of Brazil, reaching the state of Rio Grande do Sul located at the extreme meridional region of the country (Brazilian Ministry of Health, 2020a; Almeida et al., 2012; Romano et al., 2014). During these outbreaks, thousands of NHPs deaths were documented, mostly in *Alouatta* sp. (i.e. howler monkeys), followed by *Callithrix* sp. and *Cebus* sp. (Romano et al., 2014, Chame et al., 2020), and in the most recent circulation, for the first time the virus was detected in *Leontopithecus* sp., an endemic highly endangered species (Silva et al., 2020). Additionally, more than 2,100 human cases were reported in the Southeast region of Brazil, with a case-fatality rate of ∼30%, many of them in areas with low human vaccination coverage (Brazilian Ministry of Health, 2020ab).

In September 2020 a novel YFV re-emergence was observed within the midwestern states of Goiás, Distrito Federal and the Southeast state of Minas Gerais (Brazilian Ministry of Health, 2021/2022; Minas Gerais State Department of Health, 2022). This re-emergence was concomitant with the outbreak that started in 2016 and is ongoing in the Southern region of the country (Andrade et al., 2021), increasing the complexity of the Brazilian epidemiological scenario and the risk to public health (Silva et al., 2020; Brazilian Ministry of Health, 2021/2022). While the urban cycle of yellow fever has been absent in Brazil since 1942 (Possas et al., 2018), these recent outbreaks have occurred in proximity to areas heavily infested by potential *Ae. aegypti* and *Ae. albopictus* vectors that are close to large, densely populated metropolitan regions. These areas are also characterised by low YFV vaccination coverage, thereby representing a risk for the re-establishment of the urban cycle (Romano et al., 2014). Previous studies have analysed the spatial and evolutionary dynamics of the current YFV outbreak in different Southeastern states (Faria et al., 2018) and have documented the co-circulation of distinct YFV lineages (Delatorre et al., 2019; Giovanetti et al., 2019). Nevertheless, a shortage of genomic data from many locations and time points has hampered the ability to understand in detail the reemergence and establishment of YFV transmission across extra-Amazonian regions (Giovanetti et al., 2019; Faria et al., 2018).

Identifying spatial corridors of YFV spread, their ecological backgrounds, the underlying human immunity landscape, as well as the role of the animal vertebrate hosts and vector populations are crucial to predicting, preventing and controlling future potential outbreaks events that may lead to epidemics. To gain better insights into the routes of YFV dispersion, we tracked the spread and re-emergence of the virus by analysing metadata on human infections and newly 147 genome sequences sampled mainly from NHPs, as well as from some human patients within the Northern, Midwestern, Southeastern, and Southern regions of Brazil.

## Material and methods

### Sample collection

Human and non-human primate samples were collected by local and regional public health authorities, under the guidelines of a national strategy of yellow fever surveillance and sent for molecular diagnostics to the Reference Laboratory of Emerging Viruses of the Carlos Chagas Institute/Fiocruz-PR, which is a Brazilian Ministry of Health Regional Reference Laboratory for arboviruses.

### Ethical statement

The project was supported by the Pan American World Health Organization (PAHO) and the Brazilian Ministry of Health (MoH) as part of the arboviral genomic surveillance efforts within the terms of Resolution 510/2016 of CONEP (Comissão Nacional de Ética em Pesquisa, Ministério da Saúde; National Ethical Committee for Research, Ministry of Health).

### Molecular screening and nanopore sequencing

RNA was extracted from tissue samples using the QIAamp Viral RNA Mini KitTM (Qiagen, Hilden, Germany) or the MagMAX pathogen RNA/DNA kit (Life Technologies, Carlsbad, USA) according to the manufacturer’s instructions. YFV RNA was detected using the RT-qPCR protocol described by Domingo et al (Domingo et al., 2012).

### cDNA synthesis and whole-genome nanopore sequencing

Sequencing was attempted on 147 selected RT-PCR-positive samples (from a total of 200) regardless of CT value (as previously described) and the availability of epidemiological data (Faria et al., 2018; Giovanetti et al., 2019). All positive samples were submitted to a cDNA synthesis protocol (Faria et al., 2018) using a ProtoScript II first strand cDNA synthesis kit. A multiplex tiling PCR was then performed using the previously published YFV primer scheme and 30 cycles of PCR using Q5 high-fidelity DNA polymerase (NEB) as previously described (Quick et al., 2017). Amplicons were purified using AMPure XP beads (Beckman Coulter), and cleaned-up PCR product concentrations were measured using a Qubit double-stranded DNA (dsDNA) high-sensitivity (HS) assay kit on a Qubit 3.0 fluorometer (Thermo Fisher). DNA library preparation was performed using the Ligation sequencing kit (Oxford Nanopore Technologies) and the Native barcoding kit (NBD103; Oxford Nanopore Technologies, Oxford, UK). A sequencing library was generated from the barcoded products using the genomic DNA sequencing kit SQK-MAP007/SQK-LSK208 (Oxford Nanopore Technologies). The sequencing library was loaded onto a R9.4 flow cell (Oxford Nanopore Technologies). Sample data, location, hosts, NHP species, municipality of collection and collection date are summarised in Table 1.

### Generation of consensus sequences

Raw files were basecalled using Guppy v4.5.4 and barcode demultiplexing was performed using qcat. Consensus sequences were generated by *de novo* assembling using Genome Detective (https://www.genomedetective.com/) (Vilsker et al., 2019). Briefly, Genome Detective uses DIAMOND to identify and classify candidate viral reads in broad taxonomic units, using the viral subset of the Swissprot UniRef90 protein database. Candidate reads are next assigned to candidate reference sequences using NCBI blastn and aligned using AGA (Annotated Genome Aligner) and MAFFT. Final contigs and consensus sequences are made available as FASTA files. More detail about Genome Detective can be found in (Vilsker et al., 2019).

### Collation of YFV complete genome data sets

Genotyping was first conducted using the yellow fever typing tool available at https://www.genomedetective.com/app/typingtool/yellowfever/ and confirmed using a maximum likelihood (ML) phylogenetic analysis incorporating a collection of representative sequences (n= 495) belonging to the four YFV lineages (**Fig S1**). This analysis robustly placed the novel YFV sequences generated here into a clade that likely emerged in the state of Roraima in North Brazil in 2002 (**Fig S1**).

The genome sequences generated here were combined with a data set comprising previously published genomes from the 2002, 2016 to 2019 YFV epidemics in Brazil (Faria et al., 2018; Delatorre et al., 2018; Giovanetti et al., 2019). Three complete or near complete YFV genome data sets were generated. Data set 1 (n=443) comprised the data reported in this study (n=147) plus (n=296) complete or near complete YFV genomic sequences (10,000 bp) retrieved from NCBI in September 2022 and covering all the YFV South American (SA) I genotype genomes currently available. Subsequently, to understand the transmission and the spatiotemporal evolution of the SA1 lineages 1 and 2 of YFV, genetic analyses were conducted on smaller data sets that included all available genomic sequences belonging to each of those lineages: n=163 and n=270, respectively. Sequence alignment was performed using MAFFT (Katoh et al., 2013) and manually curated to remove artefacts using Aliview (Larson et al., 2014). ML phylogenetic trees were estimated using IQ-TREE2 (Nguyen et al., 2015) under the GTR nucleotide substitution model, which was inferred as the best-fit model by the ModelFinder application implemented in IQ-TREE2 (Kalyaanamoorthy et al., 2017). Statistical support for tree nodes was estimated using a ML bootstrap approach with 1,000 replicates. To investigate the temporal signal in our YFV data sets, we regressed root-to-tip genetic distances from this ML tree against sample collection dates using TempEst v.1.5.1 (Rambaut et al., 2016).

### Dated phylogenetics

To estimate time-calibrated phylogenies, we conducted a phylogenetic analysis using a Bayesian approach (Suchard et al., 2018). Accordingly, we used the GTR+gamma4 nucleotide substitution model and Bayesian Skyline tree prior as employed previously (Giovanetti et al., 2019) with an uncorrelated relaxed clock with a lognormal distribution (Giovanetti et al., 2019). Analyses were run in duplicate in BEAST v.1.10.4 for 100 million MCMC steps, sampling parameters, and trees every 10,000th step. Convergence of MCMC chains was checked using Tracer (Rambaut et al., 2018). Maximum clade credibility (MCC) trees were summarised using TreeAnnotator after discarding 10% as burn-in.

### Phylogeographic analyses

To model the phylogenetic diffusion of YFV SA1 lineages 1 and 2 we used a flexible relaxed random walk diffusion model (Lemey et al., 2010, Pybus et al., 2012) that accommodates branch-specific variation in rates of dispersal with a Cauchy distribution and a jitter window site of 0.01 (Dellicour et al., 2016, Dellicour et al., 2021). For each sequence, coordinates of latitude and longitude were attributed. MCMC analyses were performed in BEAST v1.10.4, running in duplicate for 100 million interactions and sampling every 10,000 steps in the chain. Convergence for each run was assessed in Tracer (effective sample size for all relevant model parameters >200). MCC trees for each run were summarised using TreeAnnotator after discarding the initial 10% as burn-in. Finally, we used the R package ‘seraphim’ version 1.0 (Dellicour et al., 2016) to extract and map spatiotemporal information embedded in the posterior trees.

### Eco-epidemiological data and integration with genomic data

Data of weekly notified and laboratory confirmed cases of infection by YFV in Brazil during 2015 to 2022 were supplied by the Brazilian Ministry of Health (BrMoH). A mosquito-viral suitability measure (index P) was estimated using the MVSE R-package v1.01 (Obolski et al., 2019). The index P receives as input local temperature, humidity and proposed probability distributions for key viral, vector and host parameters related to the host-pathogen system under study. Accordingly, it measures the reproductive (transmission) potential of a single adult female mosquito in a completely susceptible host population. For parameterization we used satellite climate data from Copernicus.eu (dataset “ERA5-Land monthly averaged data from 1950 to present”, https://cds.climate.copernicus.eu/), and parameter probability distributions informed by the literature. A full description of Index P parameterization can be found in the **Supplementary Text**. We used R v4.1.2 to calculate the correlation between incidence and index P using Spearman’s rank correlation coefficient (function cor.test) and to determine best time lag between the two time series (function ccf).

### Bayesian modelling of YFV human case counts

We modelled human case counts and candidate covariates by municipality using the Besag York Mollié model (BYM2). Briefly, this model assumes a Poisson distribution for case counts and includes both spatial covariates and random effects to account for unexplained spatial variation. The model was implemented in R v4.1.2 and Stan. A full description of covariates, the model and its output (posterior distributions) are provided in the **Supplementary Text**.

## Results

### Human incidence of YFV in Brazil

Due to inherent natural variations in mosquito population size, YFV should display temporal dynamics with an oscillatory behaviour characterised by recurrent peak incidence every epidemic season (local mid-summer) (Couto-Lima et al., 2017). However, limited surveillance and local testing capacity mean that obtaining a detailed spatio-temporal perspective with high resolution, and within Brazilian microregions, is challenging. Nonetheless, the epidemic curves of human case reports in the study period showed clear seasonal outbreaks within several national macroregions, compatible with the periods previously detected and used for national surveillance (Romano et al., 2011) (**Fig 1a**). In particular, between 2015 and 2022 there were three outbreaks in the Southeast (2016-2019), followed by two smaller outbreaks in the South (2020-2021). Estimated transmission potential of YFV (index P) based on local climatic variables averaged across the macroregions presented clear seasonal oscillations matching the time windows of the observed waves of infection (**Fig 1a**). The majority of human reported infections also took place in inter-yearly periods that had higher index P values (**Fig 1a inner plot**). When looking at the years 2017-2019 for which incidence presented a clearer seasonal signal, we found a positive correlation between cases and index P of 0.47 (p-value=0.003). As shown in previous studies, index P often precedes incidence in time due to inherent natural lags between natural climate variation and its effects on transmission (Nakase et al., 2022), or due to delays in reporting. Accordingly, we found that an optimal shift of index P by +1 month into the future maximised the correlation with incidence at 0.73 (p-value=5.7e-07).

**Figure 1.**
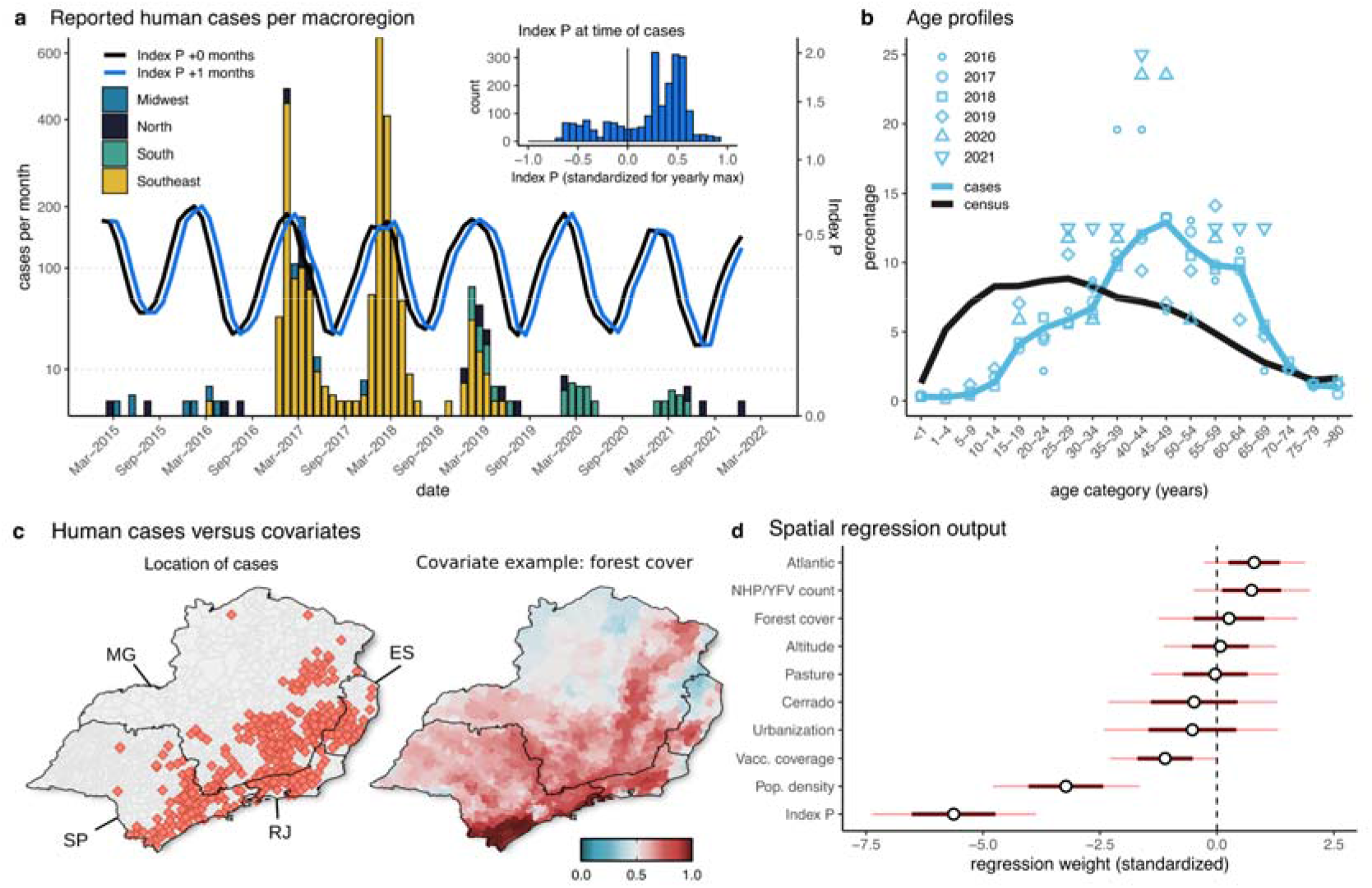
Human incidence of YFV in Brazilian macroregions, 2015–2022. **(a)** Time series of monthly reported human cases in four Brazilian macroregions (those reporting the majority of cases) and monthly mosquito-viral suitability measure (index P) aggregated (mean) across the macroregions. Index P is presented by the lines as baseline (black) and shifted by +1 month (blue). The inset plot shows the distribution of human cases according to the value of Index P (shifted +1 month) in the same month of occurrence (standardised by yearly maximum). **(b)** Age distribution of human cases across the years (light blue) versus Census 2010 data (black), both aggregated across the Southeast and South macroregions. **(c)** Spatial distribution of human cases (red filled points) and one example of a spatial covariate (forest cover) used for modelling the Southeast macroregion. For all covariates see **Fig S2**. Covariates were normalised by their maximum for visualisation, with covariates population density, Atlantic and NHP cases first transformed with log10 (colour scale in the bottom right). Spatial boundaries (black) are states: São Paulo (SP), Espírito Santo (ES), Minas Gerais (MG), Rio de Janeiro (RJ). **(d)** Marginal posterior distributions (standardised to appear on the same scale) for individual regression weights for the 10 covariates explored.

In accordance with previous studies (e.g. Tuboi et al., 2007), we found that incidence across the entire time and spatial ranges of observation were characterised by an older age profile compared to that of the population within the macroregions (**Fig 1b**). Overall, the highest burden was found in the 45-49 age group (∼13% of cases) which was at odds with a much younger age profile of the local population. Also in agreement with previous studies (Tuboi et al., 2007; Faria et al., 2018), incidence was higher among males (∼82% of all cases), in a background of ∼49% of the population being male within those macroregions.

We explored the spatio-temporal association between human case counts with ten covariates at the municipality level (**Figures S2-3**), including the suitability index P, human population density, vaccination coverage, Cerrado and Atlantic forest biomes, pasture and urbanisation land uses, forest cover and NHP YFV counts (**Tables S2-3**). We focused on the states of São Paulo, Minas Gerais, Espírito Santo and Rio de Janeiro, which together reported 2,210 of the 2,289 human cases (97%) in our data set (**Fig 1c**). To quantify the roles of each covariate we applied a Bayesian spatial regression model at the municipality level (**Supplementary Text**). Our analysis revealed considerable overdispersion due to unobserved random spatial effects (**Fig S4**), such that the ten covariates could not explain the spatial distribution of reported human cases alone. The estimated posterior distributions for the regression coefficients of each covariate (**Fig 1d, Table S4**) showed results compatible with expectations from the visual inspection of spatial distributions, with four variables having positive effects and six having negative effects (**Fig 1d, Fig S2**). Of note, there was a positive association of human cases with the Atlantic forest biome and occurrence of NHP/YFV cases, and negative associations with vaccination coverage, population density and index P (**Fig 1d**). The index P had the strongest negative effect, which contrasted the positive time-based correlation of human cases with the index (**Fig 1a**). Together, these observations revealed that while the timing of spillover events are largely dictated by climate seasonality (**Fig 1a**), the locations of spillover events are not necessarily the ones with highest suitability due to more relevant local factors such as forest biome type, forest cover and altitude that are all negatively associated with the spatial distribution of index P (**Fig S5**). This mix of covariate contributions reinforced the notion that reported human cases typically result from transmission events associated with regions reporting NHP cases within or bordering forested environments, with lower vaccine coverage and human population density (Kaul et al., 2018; Cunha et al., 2019), which are not necessarily regions with the most favourable conditions for transmission by *Aedes aegyti*.

We further mapped the predicted probability of human YFV case reporting (from the spatial regression model) and the covariate of vaccine coverage to obtain their bivariate spatial variation (**Figs 2a-b**). Spatial hotspots emerged with both high probability of reporting and proportion unvaccinated towards the southeast of the study area (**Fig 2c**). While recent past reporting was negatively associated with human population density (**Fig 1d**), many of the largest cities in the area of study were found to be peripheral to these hotspots, highlighting areas with large human populations susceptible to future outbreaks events (**Fig 2c**). This was particularly the case for the states of São Paulo (cities of São Paulo, Campinas, Guarulhos), Minas Gerais (Belo Horizonte, Contagem) and Espiríto Santo (Serra, Vila Velha, Cariacica). For the state of Rio de Janeiro, the largest cities were characterised by a mix of a high proportion of unvaccinated individuals and an intermediate probability of reporting, but were closely surrounded by hotspots of low vaccination coverage and high levels of reporting to the north. The only large city seemingly not closely connected to such a hotspot was Uberlândia (Minas Gerais), which had intermediate vaccination levels and low risk of occurrence. The identification of these hotspots, characterised by a mix of historical case reporting and a proportion of unvaccinated individuals living in close proximity to the largest cities, highlight areas of critical importance in which vaccination coverage and surveillance need to be enhanced in the near future.

**Figure 2.**
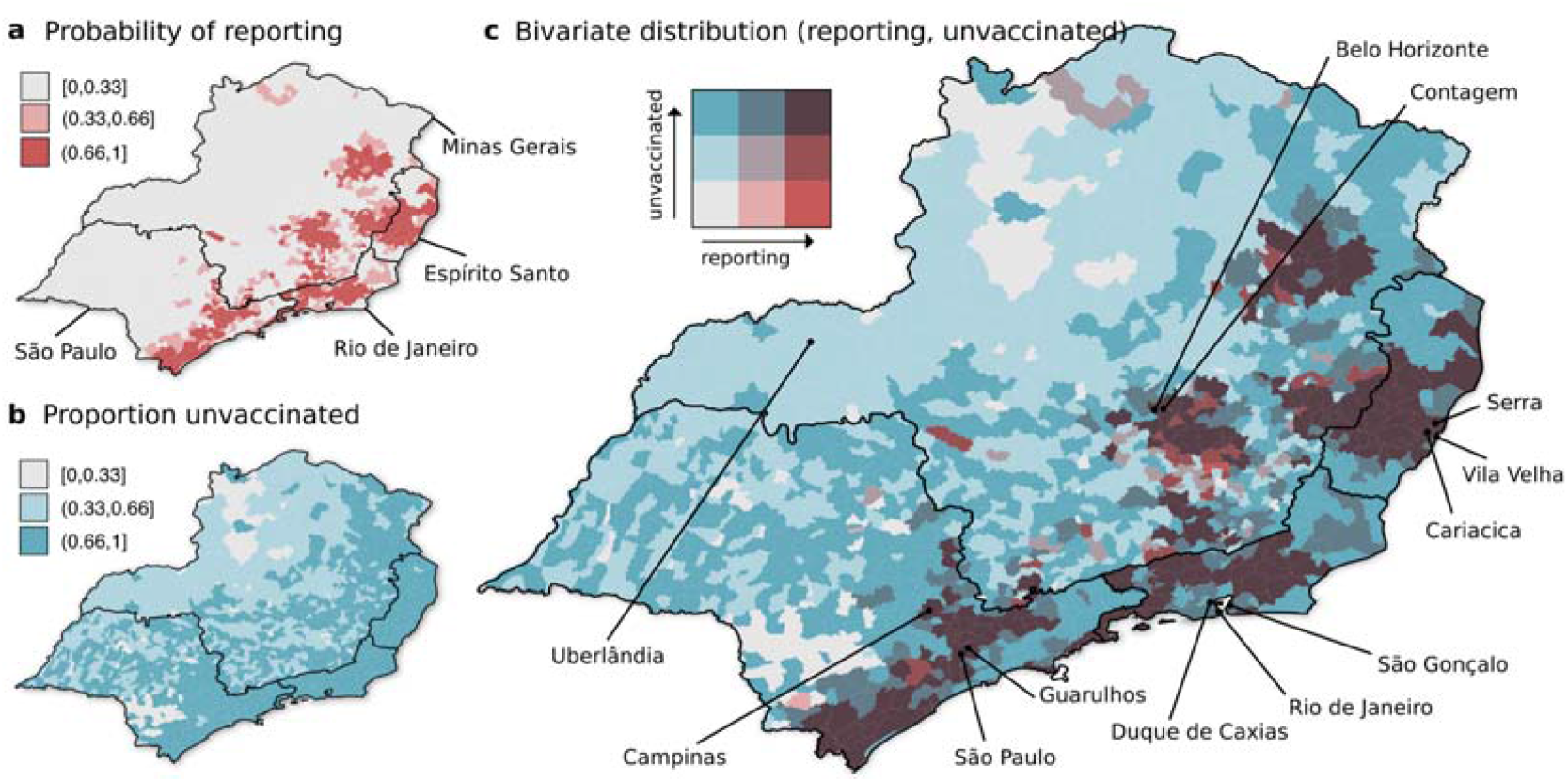
Spatial hotspots as a mixture of past reporting and proportion unvaccinated. **(a)** Probability of reporting of human YFV cases per municipality as predicted by the spatial regression model. **(b)** Proportion unvaccinated (1 - vaccination coverage) per municipality. **(a-b)** Variables are discretized into three categories to provide a bivariate colour scale in panel c with a maximum of nine colours for best interpretability. **(c)** Spatial distribution of both probability of occurrence and proportion unvaccinated presented in bivariate colour scale. The three largest cities in each state are highlighted (São Paulo: São Paulo, Guarulhos, Campinas; Minas Gerais: Belo Horizonte, Uberlândia, Contagem; Rio de Janeiro: Duque de Caxias, Rio de Janeiro, São Gonçalo; Espírito Santo: Serra, Vila Velha, Cariacica).

### Phylodynamics of YFV in Brazil

To explore the phylodynamics of YFV in Brazil, we combined our 147 newly generated sequences obtained from the Southeastern, Midwestern and Southern macroregions of the country to those of other genomes available on GenBank (n=296). Our analysis revealed the circulation of three different clades, named hereafter as clades Ia, IIb, as well as a novel clade termed IIIc (**Fig 3**). As previously described (Delatorre et al., 2019; Giovanetti et al., 2019), the South American genotype I was characterised by the co-circulation of two viral lineages (i.e., clades Ia and IIb). Circulating clades Ia-IIIc were characterised by specific mutational signatures, including nonsynonymous changes in the RNA-dependent-RNA polymerase (RdRp): S8053D and T9806I in clade Ia, V8048I in clade IIb, and G8048A in clade IIIc. These four nonsynonymous and other six synonymous mutations are described in **Table S5**.

**Figure 3.**
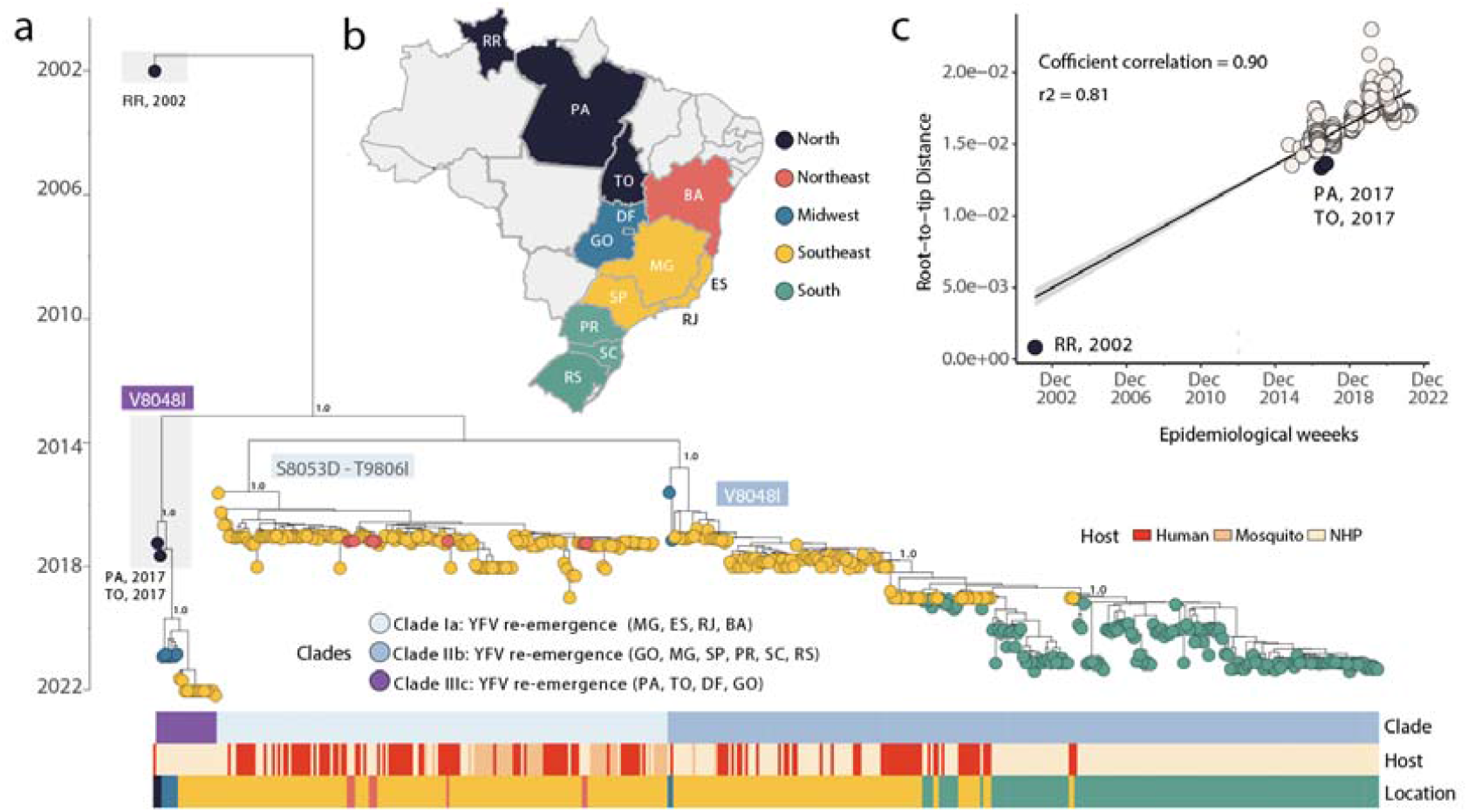
YFV South America I genotype in Brazil. **(a)** MCC phylogeny inferred using the 147 novel sequences obtained in this study plus 296 publicly available sequences from GenBank. Colours represent different sampling locations (Brazilian states). Colored bars below the tree represent the (top) clade (middle) host and Brazilian macroregion of sampling. **(b)** Map of Brazil presenting the states under investigation highlighted within macroregions: RR= Roraima, PA= Pará, DF = Distrito Federal, GO= Goiás, BA = Bahia, MG= Minas Gerais, ES= Espírito Santo, RJ= Rio de Janeiro, SP= São Paulo state, PR = Paraná, SC= Santa Catarina, RS= Rio Grande do Sul. (**c)** Root-to-tip regression of sequence sampling date against genetic divergence from the root of the outbreak clade. Sequences from the endemic Amazon basin (states of Roraima, Pará and Tocantins) are highlighted.

To investigate the evolution of clades Ia and IIb in more detail, we used smaller data sets derived from each clade individually. Before phylogeographic analysis, each clade was also assessed for molecular clock signal using the root-to-tip regression method available in TempEst v1.5.3 (Rambaut et al., 2016), following the removal of potential outliers that may strongly violate the assumption of rate constancy. The final data set included n= 163 genome sequences for Clade Ia, and n= 270 genome sequences for Clade IIb. Importantly, there was a strong correlation between sampling time and the root-to-tip divergence in the two data sets (**Figs 4a, 5a**), indicative of a constancy of evolutionary rate and hence allowing the use of molecular clock models to infer evolutionary parameters. Our phylogeographic reconstruction of clade Ia (**Fig 4b**) suggested a mean time of origin in late-July 2015 (95% highest posterior density (HPD): 29 March 2015 to 18 August 2015). Viruses from this clade spread from the Northern region of the state of Minas Gerais towards the Southeast and later to the Northeast, as indicated by isolates from the Bahia state.

**Figure 4.**
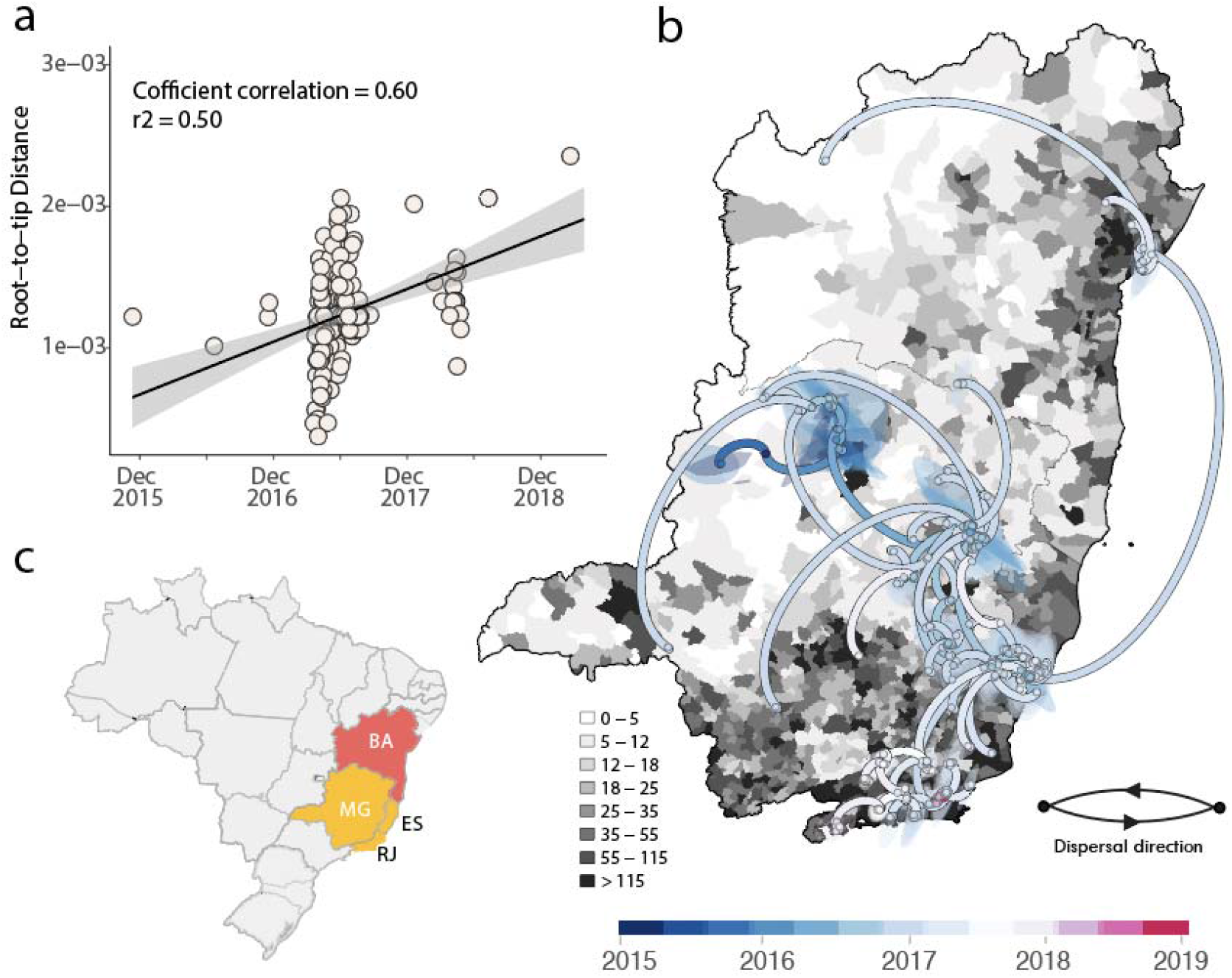
Spatiotemporal spread of South America I genotype Clade Ia in Brazil. a) Root-to-tip regression of sequence sampling date against genetic divergence from the root of the outbreak clade; b) Phylogeographic reconstruction of the spread of the South America I genotype Clade Ia in Brazil (n=163). Circles represent nodes of the MCC phylogeny and are coloured according to their inferred time of occurrence. Shaded areas represent the 80% highest posterior density interval and depict the uncertainty of the phylogeographic estimates for each node. Solid curved lines denote the links between nodes and the directionality of movement. Differences in population density are shown on a grey-white scale. c) Map of Brazil highlighting the spatial area under investigation: MG= Minas Gerais state, ES= Espírito Santo state, RJ= Rio de Janeiro state, BA= Bahia state.

**Figure 5.**
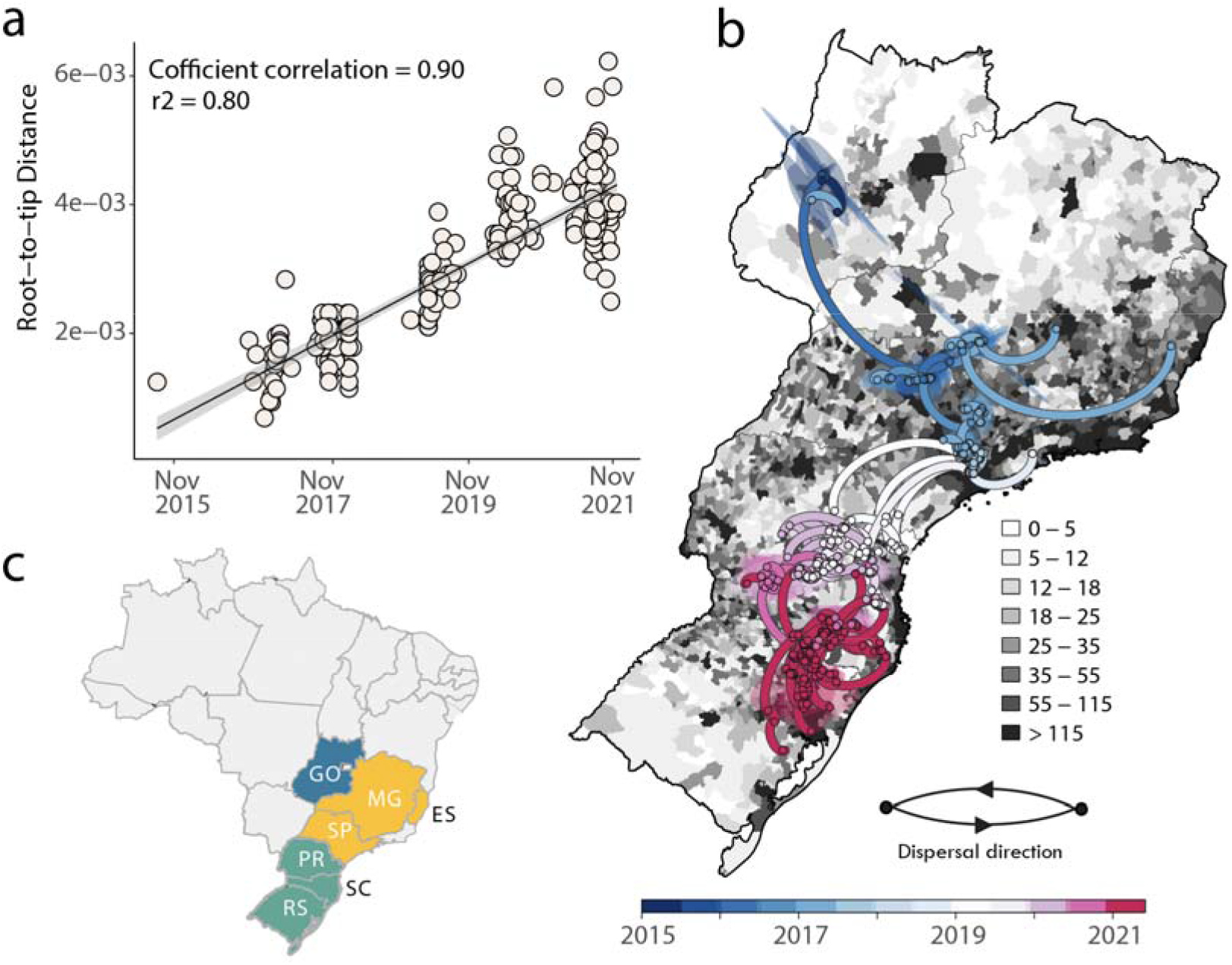
Spatiotemporal spread of South America I genotype Clade IIb in Brazil. a) Root-to-tip regression of sequence sampling date against genetic divergence from the root of the outbreak clade. b) Phylogeographic reconstruction of the spread of South America I genotype Clade IIb in Brazil (n=270). Circles represent nodes of the MCC phylogeny and are coloured according to their inferred time of occurrence. Shaded areas represent the 80% highest posterior density interval and depict the uncertainty of the phylogeographic estimates for each node. Solid curved lines denote the links between nodes and the directionality of movement.Differences in population density are shown on a grey-white scale. c) Map of Brazil highlighting the spatial area under investigation: GO= Goiás state, MG= Minas Gerais state, ES= Espírito Santo state, RJ= Rio de Janeiro state, SP= São Paulo state, PR= Paraná state, SC= Santa Catarina state, RS= Rio Grande do Sul state.

A different spatial pattern was observed for clade IIb (**Fig 5**) compared to clade Ia (**Fig 4**). Specifically, clade IIb originated with a mean date in late November 2015 (95% highest posterior density (HPD): 28 November 2015 to 05 January 2016) with an early dispersal from the Midwest (state of Goiás) to the Southeast (Minas Gerais, Espírito Santo and São Paulo) and later to the South (state of Paraná, Santa Catarina and Rio Grande do Sul) where it persisted at least until the end of 2021 (**Fig 5b**).

Through analysis of the genomic data set that included samples collected in 2017 from the North macroregion we were able to identify a corridor of viral spread associated with clade IIIc (Figure 3, posterior probability - pp 1.0) that connected the endemic North with the extra-Amazonian basin (states of Goiás and Distrito Federal in the Midwest, and the state of Minas Gerais in the Southeast). We estimated the mean time of the most recent common ancestor (tMRCA) for this clade to be July 2016 (95% highest posterior density interval [HPD], April 2015 - March 2017).

## Conclusion / discussion

The circulation of YFV has been reported in the extra-Amazonian regions of Brazil since the early 2000s (Brazilian Ministry of Health, 2020a; Almeida et al., 2012; Romano et al., 2014; Vasconcelos et al., 2003; Cardoso et al., 2010). During this period, many advances have been achieved in the country regarding surveillance in both the reservoir and human population, such as the start of many digital platforms for animal and case reporting (e.g. https://sissgeo.lncc.br/). Preliminary studies demonstrated that a later reemergence, which began in 2016, resulted in widespread virus dissemination and an extended period of transmission still active in 2022 (Andrade et al., 2021, Andrade et al., 2022). The uninterrupted circulation of YFV in the extra-

Amazonian environment for a period that exceeds seven years is unprecedented and coincides with ongoing ecological changes. For example, increases in temperature and precipitation in some areas of the Atlantic forest biome in the South and Southeast may be contributing to creating conditions for the maintenance of perennial foci of YFV transmission in or around highly populated areas (Marengo et al., 2020; Regoto et al., 2021). At the same time, while it remains difficult to quantify direct causal effects of landscape changes due to human intervention, the midwestern and southeastern states analysed in this study are known to be experiencing rapid alterations. Indeed, changes in the forestry legislation that took effect in 2016 have allowed and fomented degradation of forested environments; for example decreasing the size of protected forests along rivers that are expected to increase exposure of adult, male and poor rural workers involved in logging and farming (Dobson et al., 2020; Ribeiro et al., 2022). By 2017, a record primary forest deforestation was measured along the Doce river valley in the state of Minas Gerais (Global forest, 2022). Similarly, the number of forest fires in the region where YFV re-emerged in 2020 more than doubled in 2018 and 2019 (National Institute of Spatial Research, 2022). Such landscape changes are driven by the need to de-forest environments for agricultural purposes on a large scale, and 2020 may represent a year of greater exposure of workers to degraded forests and secondary forests (Brondízio et al., 2009; Schneider et al., 2015). This recent trend is unlikely to change drastically in coming years, and the exploration of the role of degraded forested environments in increased risk of YFV spillover from the non-human primate hosts should be the focus of future research.

In recent years, thousands of cases and deaths, mainly among NHPs, have caused unprecedented impacts on these animal populations and public health, resulting in raised awareness and control activities, such as the increase of vaccine coverage to classically non-endemic territories (Andrade et al., 2021, Andrade et al., 2022). These recent trends also relate to conservation efforts for NHPs. For example, in the context of the endangered *Alouatta guariba clamitans*, previous work suggests that YFV infection is a major threat for population stability, since outbreaks in fragmented, metapopulation-structured populations are likely to result in local extinctions (Moreno et al., 2015, Chame et al. 2020). In parallel, an ongoing decline in the size of the Atlantic forest protected areas, due to recent government policies and the abandonment of surveillance and law enforcement, might have both increased the risks of new NHP outbreaks and, consequently, chances the infection to humans in the environs of areas with high population density.

Previous studies have examined the spatial and evolutionary dynamics of YFV between 2016 and 2019 (Faria et al., 2018; Giovanetti et al., 2019). Unfortunately, however, the shortage of genomic data has hampered our ability to understand particular aspects of this epidemic, such as the current reemergence in the Southeast (since late 2019). In this study, we presented a combination of epidemiological and genomic analysis of novel YFV data with the aim of further understanding and describing the past and present of the ongoing epidemic.

A time series of reported human cases between 2015 and 2022 showed a typical yearly seasonal pattern associated with a midsummer peak in January, as well as three outbreaks in the Southeast (2017-2019) followed by two outbreaks in the South (2020-2021). In general, reporting was associated with areas peripheral or within forested environments characterised by an Atlantic forest biome, to males and older age groups, to variation in climate, population density and vaccination coverage. The suitability index used was calibrated to the urban cycle vector *Aedes aegypti* due to lack of data for mosquito-vectors of the enzootic transmission cycle. The index was associated positively with the timing of reported human infections, providing opportunities to estimate time-windows of importance for spillover events and surveillance across the country in future studies.

We were able to identify spatial hotspots characterised by both the reporting of human cases and low vaccination coverage, which were critically peripheral to some of the largest human populations in the Southeast. These hotspots demonstrate the recent reporting of cases close to large cities with large susceptible human populations and where *Aedes aegypti* is also typically abundant, thereby presenting possible ecological boundaries across which urban transmission cycles could become established. A lack of access to detailed reporting of human cases in other regions of Brazil hampered the application of our spatial regression model to a wider spatial scale. However, it remains plausible that similar hotspots exist within other states, including the state of Bahia that served as a corridor for clade Ia into the southern states, is known to be rich in NHP biodiversity, and also harbours large human populations. These hotspots in the Southeast should become the focus of future targeted mosquito and NHP surveillance, as well as public health campaigns to boost public awareness and vaccination coverage in particular within the peripheries of large urban centres.

Our newly generated YFV sequences were classified to the South American I genotype and formed three distinct clades Ia–IIIc. These clades presented different spatial patterns. Clade Ia historically spread from the Northern region of the state of Minas Gerais towards the Southeast and later to the Northeast. In contrast clade IIb showed an early dispersal from the Midwest (state of Goiás) to the Southeast (Minas Gerais, Espírito Santo and São Paulo) with a later spread to the South (state of Paraná, Santa Catarina and Rio Grande do Sul) where it has persisted at least until the end of 2021. While the general spatial spread of clade Ia and IIb have already been described in the Brazilian context (Delatorre et al., 2018; Giovanetti et al., 2019), the analysis of the novel isolates from three macro regions in 2020 and 2021 is novel (North, Midwest and Southeast). This also allowed us to produce the first genetic evidence in support of a corridor of spread (previously described using NHP case count data (Possas et al., 2018)) associated with clade IIIc which connected the endemic North with the extra-Amazonian basin (states of Goiás, Distrito Federal in the Midwest, and the state of Minas Gerais in the Southeast). We additionally described several point mutations within clades I-III viral genomes, of which six were associated with amino acid substitutions: S8053D and T9806I in the RdRp gene of clade Ia, V8048I in the RdRp gene of clade IIb, and S8048N in the RdRp gene of clade IIIc. As the RdRp is responsible for the replication of the viral genome, further studies are required to elucidate the possible impact of these mutations on structure and function, and hence on both viral pathogenesis and fitness.

We failed to identify if the locations within this corridor (that cuts through the centre of the country) had ecological or landscape features of relevance. For example, the associated central regions of Brazil are rich in the Cerrado biome (Tisler et al., 2022) which was negatively associated with human YFV case reporting. Compared to other regions, these regions are also not particularly suitable for the *Haemagogus and Sabethes* spp. vector-species that are associated with the sylvatic cycle (Li et al., 2022), nor do they present particular richness of relevant NHP species (IUCN. 2022). A potential contributing factor could be the large rivers that stem in a north-south axis over the corridor (e.g. Rio Tocantins, Rio das Mortes, Rio Araguaia), from the state of Pará crossing Tocantins (North) into Goiás (Midwest), the latter at the north border of Minas Gerais (Southeast) as well as the forests on the edges of the north-south oriented mountains that make up the brazilian central plateau. Rivers, mountains and their associated forest environments offer ideal corridors for NHP settlement and migration, potentially contributing to the spatial spread of YFV (Dietz et al., 2019). In this study we present the first evidence of the north-south directionality of spread over this spatial corridor, which appears particularly related to degraded and deforested riparian forests, raising awareness to its public health importance and suggesting that future research and surveillance initiatives should focus on the associated regions.

Our results reinforce the importance of the North of Brazil as a potential hotspot and revealed a new role for the Midwest regions of the country, in line with studies that have highlighted both regions as relevant hubs, not only for YFV, but also for dengue virus (DENV) (Faria et al., 2018; Giovanetti et al., 2019; Adelino et al., 2021). There were critical gaps in existing genomic data (445 genomes since 2002 with patchy temporal and spatial sampling, **Fig S6**) which curtailed definite conclusions in this study on key points of the recent history of YFV in Brazil. There remains, for instance, only a small number of viral genomes from the North, which is a key region for understanding YFV persistence dynamics and genetic diversity. At the same time, although the virus continuously expands its geographical range, there has been historically weak sampling in some states such as Goiás, Distrito Federal and Minas Gerais that hampered the unravelling of the corridor of spread revealed in the current study.

By identifying spatial corridors of spread, their eco-epidemiological backgrounds and drivers and mutational signatures associated with successful viral lineages, the combination of epidemiological and genomic surveillance within a One Health approach can have significant public health impact. It can identify the likely places for the reemergence of urban cycles of transmission, inform on emerging viral lineages that should be the focus of empirical laboratory experimentation, and critically identify areas and time windows where catch-up vaccination campaigns and surveillance initiatives should be directed. Given the existing large gaps in knowledge, there is clearly a need for continued funding for genomic surveillance of YFV both in Brazil and globally.

## Data Availability

New sequences generated as part of this study have been deposited in GenBank under accession numbers OP508570-OP508716 (Table 1). Aggregate estimates of YFV temporal suitability (index P) and its input (humidity, temperature, rainfall) for the South and Southeast between 2014 and 2021 are provided as a comma separated values (CSV) file (Supplementary Data File 1). Estimates of the probability of reporting human cases per municipality for the South and Southeast (output of spatial regression model) are provided as a comma separated values (CSV) file (Supplementary Data File 2) geocoded with GEOCMU7 (a seven digit unique identifier).

## Data availability

New sequences generated as part of this study have been deposited in GenBank under accession numbers OP508570-OP508716 (**Table 1**). Aggregate estimates of YFV temporal suitability (index P) and its input (humidity, temperature, rainfall) for the South and Southeast between 2014 and 2021 are provided as a comma separated values (CSV) file (**Supplementary Data File 1**). Estimates of the probability of reporting human cases per municipality for the South and Southeast (output of spatial regression model) are provided as a comma separated values (CSV) file (**Supplementary Data File 2**) geocoded with GEOCMU7 (a seven digit unique identifier).

## Supplementary files

**Supplementary Text File 1** includes a full description of the modelling approach to YFV suitability (Index P) and spatial regression of human infections (BYM2), details on the mutational differences between YFV clades I-III, as well as Supplementary Tables S1-5 and Supplementary Figures S1-4.

## Acknowledgments

This work was supported in part through National Institutes of Health USA grant U01 AI151698 for the United World Arbovirus Research Network (UWARN) and the Brazilian Ministry of Health (SCON2021-00180). MG is funded by PON “Ricerca e Innovazione” 2014-2020. JL is funded by BioISI (Biosystems and Integrative Sciences Institute), Faculdade de Ciências da Universidade de Lisboa (UIDP/4046/2020). FP is funded by the One Health Poultry Hub, a Global Challenges Research Fund (GCRF) and United Kingdom Research and Innovation (UKRI) initiative. ECH is funded by an Australian Research Council Australian Laureate Fellowship (FL170100022). CNDS was founded by CNPq (307176-2018-5). Authors additionally thanks Fiocruz/CVSLR for the logistic support.

Conceptualization: MG, CZ, JL, LCJA and CNDS

Methodology: MG, FP, CZ, VF, TN, ACK, MT, GS, GGD, AEMLM, RS, TERA, JX, CO, SPS, NRG, HF, MAMG, FL and JL.

Investigation: MG, FP, CZ, VF, TN, ACK, MT, GS, GGD, AEMLM, RS, TARA, JX, CO, SPS, NRG, HF, MAMG, FL, PHR, VLS, LAP, AFM, ILM, DERS, GRTC, MBC, FCMI, MAP, KRLJC, ARRF, CFCA, EMM, MPDA, RCR, AASC, AP, MC, LA, INR, SPR, AIB, TO, CF, NFOM, AF, CDSR, CCS, MABA, ES, JC, DAA, EK, LFM, RRG, SFC, JABF, MGDL, ILB, GMPS, MRFS, MMSC, JCSS, ABJ, EGP, LST, DAA, RM, JSR, PCLS, ASAFS, SS, GC, RS, WN, LAM, AGAF, GGP, BTDN, DBAM, ACRC, RVC, WVV, AMBF, MA, ECH, DGR, AR, JL, LCJA, CNDS.

Sampling: MG, FP, CZ, VF, TN, ACK, MT, GS, GGD, AEMLM, RS, TARA, JX, CO, SPS, NRG, HF, MAMG, FL, PHR, VLS, LAP, AFM, ILM, DERS, GRTC, MBC, FCMI, MAP, KRLJC, ARRF, CFCA, EMM, MPDA, RCR, AASC, AP, MC, LA, INR, SPR, AIB, TO, CF, NFOM, AF, CDSR, CCS, MABA, ES, JC, DAA, EK, LFM, RRG, SFC, JABF, MGDL, ILB, GMPS, MRFS, MMSC, JCSS, ABJ, EGP, LST, DAA, RM, JSR, PCLS, ASAFS, SS, GC, RS, WN, LAM, AGAF, GGP, BTDN, DBAM, ACRC, RVC, WVV, AMBF, MA, ECH, DGR, AR, JL, LCJA, CNDS.

Sequencing: MG, VF, TERA, JX, SPS, CO and NRG.

Visualization: MG, FP, VG, TN and JL.

Funding acquisition: LCJA and CNDS.

Project administration: LCJA and CNDS.

Supervision: LCJA and CNDS.

Writing – original draft: MG, CZ, FP, JL, and CNDS.

Writing – review &amp; editing: MG, FP, CZ, VF, TN, ACK, MT, GS, GGD, AEMLM, RS, TARA, JX, CO, SPS, NRG, HF, MAMG, FL, PHR, VLS, LAP, AFM, ILM, DERS, GRTC, MBC, FCMI, MAP, KRLJC, ARRF, CFCA, EMM, MPDA, RCR, AASC, AP, MC, LA, INR, SPR, AIB, TO, CF, NFOM, AF, CDSR, CCS, MABA, ES, JC, DAA, EK, LFM, RRG, SFC, JABF, MGDL, ILB, GMPS, MRFS, MMSC, JCSS, ABJ, EGP, LST, DAA, RM, JSR, PCLS, ASAFS, SS, GC, RS, WN, LAM, AGAF, GGP, BTDN, DBAM, ACRC, RVC, WVV, AMBF, MA, ECH, DGR, AR, JL, LCJA, CNDS.

## References

Adelino TÉR, Giovanetti M, Fonseca V, Xavier J, de Abreu ÁS, do Nascimento VA, Demarchi LHF, Oliveira MAA, da Silva VL, de Mello ALES, Cunha GM, Santos RH, de Oliveira EC, Júnior JAC, de Melo Iani FC, de Filippis AMB, de Abreu AL, de Jesus R, de Albuquerque CFC, Rico JM, do Carmo Said RF, Silva JA, de Moura NFO, Leite P, Frutuoso LCV, Haddad SK, Martínez A, Barreto FK, Vazquez CC, da Cunha RV, Araújo ELL, de Oliveira Tosta SF, de Araújo Fabri A, Chalhoub FLL, da Silva Lemos P, de Bruycker-Nogueira F, de Castro Lichs GG, Zardin MCSU, Segovia FMC, Gonçalves CCM, Grillo ZDCF, Slavov SN, Pereira LA, Mendonça AF, Pereira FM, de Magalhães JJF, Dos Santos Júnior ACM, de Lima MM, Nogueira RMR, Góes-Neto A, de Carvalho Azevedo VA, Ramalho DB, Oliveira WK, Macario EM, de Medeiros AC, Pimentel V; Latin American Genomic Surveillance Arboviral Network, Holmes EC, de Oliveira T, Lourenço J, Alcantara LCJ. Field and classroom initiatives for portable sequence-based monitoring of dengue virus in Brazil. Nat Commun. 2021 Apr 16;12(1):2296. doi: 10.1038/s41467-021-22607-0. PMID: 33863880; PMCID: PMC8052316.

Almeida, M.A.B. de, E. Dos Santos, J. da C. Cardoso, D.F. da Fonseca, C.A. Noll, V.R. Silveira, A.Y. Maeda, R.P. de Souza, C. Kanamura, and R.A. Brasil. 2012. Yellow fever outbreak affecting Alouatta populations in southern Brazil (Rio Grande do Sul State), 2008-2009. Am J Primatol. 74:68–76. doi:10.1002/ajp.21010.

Andrade MS, Campos FS, Campos AAS, Abreu FVS, Melo FL, Sevá ADP, Cardoso JDC, Dos Santos E, Born LC, Silva CMDD, Müller NFD, Oliveira CH, Silva AJJD, Simonini-Teixeira D, Bernal-Valle S, Mares-Guia MAMM, Albuquerque GR, Romano APM, Franco AC, Ribeiro BM, Roehe PM, Almeida MAB. Real-Time Genomic Surveillance during the 2021 Re-Emergence of the Yellow Fever Virus in Rio Grande do Sul State, Brazil. Viruses. 2021 Oct 1;13(10):1976. doi: 10.3390/v13101976. PMID: 34696408; PMCID: PMC8539658.

Andrade, Miguel & Campos, Fabrício & Oliveira, Cirilo & Oliveira, Ramon & Campos, Aline & Almeida, Marco Antônio & Fonseca, Vagner & Teixeira, Danilo & Sevá, Anaiá & Temponi, Andrea & Magalhães, Fernando & Chaves, Danielle & Pereira, Maira & Lamounier, Ludmila & Menezes, Givaldo & Aquino-Teixeira, Sandy & Santos, Maria & Bernal Valle, Sofia & Müller, Nicolas & Abreu, Filipe. (2022). Fast surveillance response reveals the introduction of a new Yellow Fever Virus sub-lineage in 2021, in Minas Gerais, Brazil. Memórias do Instituto Oswaldo Cruz. 117. 1–18. 10.1590/0074-02760220127.

Brazilian Ministry of Health. 2020a. Febre Amarela. Avaialble in: https://www.gov.br/saude/pt-br/assuntos/saude-de-a-a-z/f/febre-amarela-1.

Brazilian Ministry of Health. 2020b. Situação epidemiológica da febre amarela no monitoramento 2019/2020. Brasília, Brazil. Avaialble in: https://www.gov.br/saude/pt-br/centrais-de-conteudo/publicacoes/boletins/boletins-epidemiologicos/edicoes/2020/boletim-epidemiologico-vol-51-no-01/@@download/file/boletim-epidemiologico-svs-01.pdf.

Brazilian Ministry of Health. 2021/2022. Situação epidemiológica da febre amarela no monitoramento 2020/2021. Brasília, Brazil. Avaialble in: https://www.gov.br/saude/pt-br/centrais-de-conteudo/publicacoes/boletins/boletins-epidemiologicos/edicoes/2021/boletim_epidemiologico_svs_31.pdf.

Brondízio ES, Cak A, Caldas M, Memna CF, Bilsborrow RE, Futemma C, Ludwigs T, Moran E, Batistella M. 2009. Small farmers and deforestation in amazônia. Geophysical Monograph Series 186: 117–143.

Cardoso JC, Almeida MAB, Santos E, Fonseca DF, Sallum MAM, Noll CA, et al. Yellow fever virus in Haemagogus leucocelaenus and Aedes serratus mosquitoes, Southern Brazil, 2008. Emerg Infect Dis. 2010;16(12):1918–24.

Chame M, Abdalla L, Pinter A, Romano PMA, Krempser E, Ramos G, Passos PHO, Silva PCL, Silva GMP, Gatti RR, Augusto DA, Sianto L. Primates in SISS-Geo: Potential contributions of mobile technology, health surveillance and citizen science to support species conservation in Brazil. Neotropical primates, v. 26, p. 80–89, 2020.

Couto-Lima, D., Madec, Y., Bersot, M.I. et al. Potential risk of re-emergence of urban transmission of Yellow Fever virus in Brazil facilitated by competent Aedes populations. Sci Rep 7, 4848 (2017). https://doi.org/10.1038/s41598-017-05186-3

Cunha, M.d.P., Duarte-Neto, A.N., Pour, S.Z. et al. Origin of the São Paulo Yellow Fever epidemic of 2017–2018 revealed through molecular epidemiological analysis of fatal cases. Sci Rep 9, 20418 (2019). https://doi.org/10.1038/s41598-019-56650-1

De Souza, R.P., P.G. Foster, M.A.M. Sallum, T.L.M. Coimbra, A.Y. Maeda, V.R. Silveira, E.S. Moreno, F.G. Da Silva, I.M. Rocco, I.B. Ferreira, A. Suzuki, F.M. Oshiro, S.M.C.N. Petrella, L.E. Pereira, G. Katz, C.H. Tengan, M.M. Siciliano, and C.L.S. Dos Santos. 2010. Detection of a new yellow fever virus lineage within the south american genotype I in Brazil. J. Med. Virol. 82:175–185. doi:10.1002/jmv.21606.

Delatorre, E., F.V. Santos De Abreu, I.P. Ribeiro, M.M. Gómez, A.A. Cunha Dos Santos, A. Ferreira-De-Brito, M.S. Alberto Santos Neves, I. Bonelly, R.M. De Miranda, N.D. Furtado, L.M. Souza Raphael, L. de F. Fernandes Da Silva, M.G. De Castro, D.G. Ramos, A.P. Martins Romano, E.G. Kallás, A.C. Paulo Vicente, G. Bello, R. Lourenço-De-Oliveira, and M. Cristina Bonaldo. 2019. Distinct YFV Lineages Co-circulated in the Central-Western and Southeastern Brazilian Regions from 2015 to 2018. Front. Microbiol. 10:z1–12. doi:10.3389/fmicb.2019.01079.

Dellicour, S. et al. Relax, keep walking -a practical guide to continuous phylogeographic inference with BEAST. Mol. Biol. Evol. 38,3486–3493 (2021).

Dellicour, S., Rose, R., Faria, N. R., Lemey, P. & Pybus, O. G. SERAPHIM:studying environmental rasters and phylogenetically informed movements. Bioinformatics 32, 3204–3206 (2016).

Dietz JM, Hankerson SJ, Alexandre BR, Henry MD, Martins AF, Ferraz LP, Ruiz-Miranda CR. Yellow fever in Brazil threatens successful recovery of endangered golden lion tamarins. Sci Rep. 2019 Sep 10;9(1):12926. doi: 10.1038/s41598-019-49199-6. PMID: 31506447; PMCID: PMC6736970.

Dobson AP, Pimm SL, Hannah L, Kaufman L, Ahumada JA, Ando AW, Bernstein A, Busch J, Daszak P, Engelmann J, Kinnaird MF, Li BV, Loch-Temzelides T, Loveloy T, Nowak K, Roehrdanz PR, Vale MM. Science 369: 379–381. https://www.science.org/doi/10.1126/science.abc3189

Domingo C, Patel P, Yillah J, Weidmann M, Méndez JA, Nakouné ER, Niedrig M. 2012. Advanced yellow fever virus genome detection in point-of-care facilities and reference laboratories. J Clin Microbiol 50:4054–4060. https://doi.org/10.1128/JCM.01799-12.

Faria NR, Kraemer MUG, Hill SC, Goes de Jesus J, Aguiar RS, Iani FCM, Xavier J, Quick J, Du Plessis L, Dellicour S, Thézé J, Carvalho RDO, Baele G, Wu CH, Silveira PP, Arruda MB, Pereira MA, Pereira GC, Lourenço J, Obolski U, Abade L, Vasylyeva TI, Giovanetti M, Yi D, Weiss DJ, Wint GRW, Shearer FM, Funk S, Nikolay B, Fonseca V, Adelino TER, Oliveira MAA, Silva MVF, Sacchetto L, Figueiredo PO, Rezende IM, Mello EM, Said RFC, Santos DA, Ferraz ML, Brito MG, Santana LF, Menezes MT, Brindeiro RM, Tanuri A, Dos Santos FCP, Cunha MS, Nogueira JS, Rocco IM, et al. 2018. Genomic and epidemiological monitoring of yellow fever virus transmission potential. Science 361:894–899. https://doi.org/10.1126/science.aat7115.

Giovanetti M, de Mendonça MCL, Fonseca V, Mares-Guia MA, Fabri A, Xavier J, de Jesus JG, Gräf T, Dos Santos Rodrigues CD, Dos Santos CC, Sampaio SA, Chalhoub FLL, de Bruycker Nogueira F, Theze J, Romano APM, Ramos DG, de Abreu AL, Oliveira WK, do Carmo Said RF, de Alburque CFC, de Oliveira T, Fernandes CA, Aguiar SF, Chieppe A, Sequeira PC, Faria NR, Cunha RV, Alcantara LCJ, de Filippis AMB. Yellow Fever Virus Reemergence and Spread in Southeast Brazil, 2016-2019. J Virol. 2019 Dec 12;94(1):e01623–19. doi: 10.1128/JVI.01623-19. Erratum in: J Virol. 2020 May 18;94(11): PMID: 31597773; PMCID: PMC6912119.

Giovanetti, M., M.C.L. de Mendonça, V. Fonseca, M.A. Mares-Guia, A. Fabri, J. Xavier, J.G. de Jesus, T. Gräf, C.D. dos Santos Rodrigues, C.C. dos Santos, S.A. Sampaio, F.L.L. Chalhoub, F. de Bruycker Nogueira, J. Theze, A.P.M. Romano, D.G. Ramos, A.L. de Abreu, W.K. Oliveira, R.F. do Carmo Said, C.F.C. de Alburque, T. de Oliveira, C.A. Fernandes, S.F. Aguiar, A. Chieppe, P.C. Sequeira, N.R. Faria, R.V. Cunha, L.C.J. Alcantara, and A.M.B. de Filippis. 2019. Yellow Fever Virus Reemergence and Spread in Southeast Brazil, 2016–2019. J. Virol. 94. doi:10.1128/jvi.01623-19.

Global Forest, 2022. Report available at: https://www.globalforestwatch.org/map/.

IUCN.2022. The IUCN Red List of Threatened Species. Version 2022-1. https://www.iucnredlist.org.

Kalyaanamoorthy, S., Minh, B. Q., Wong, T. K. F., von Haeseler, A. & Jermiin, L. S. ModelFinder: fast model selection for accurate phylogenetic estimates. Nat. Methods 14, 587–589 (2017).

Katoh, K. & Standley, D. M. MAFFT Multiple Sequence Alignment Software Version 7: improvements in performance and usability. Mol. Biol. Evol. 30, 772–780 (2013).

Kaul RB, Evans MV, Murdock CC, Drake JM. Spatio-temporal spillover risk of yellow fever in Brazil. Parasit Vectors. 2018 Aug 29;11(1):488. doi: 10.1186/s13071-018-3063-6. PMID: 30157908; PMCID: PMC6116573.

Laarson, A. AliView: a fast and lightweight alignment viewer and editor for large data sets. Bioinformatics 30, 3276–3278 (2014).

Lemey P, Rambaut A, Welch JJ, Suchard MA. 2010. Phylogeography takes a relaxed random walk in continuous space and time. Mol Biol Evol 27:1877–1885. https://doi.org/10.1093/molbev/msq067.

Li SL, Acosta AL, Hill SC, Brady OJ, de Almeida MAB, Cardoso JDC, Hamlet A, Mucci LF, Telles de Deus J, Iani FCM, Alexander NS, Wint GRW, Pybus OG, Kraemer MUG, Faria NR, Messina JP. Mapping environmental suitability of Haemagogus and Sabethes spp. mosquitoes to understand sylvatic transmission risk of yellow fever virus in Brazil. PLoS Negl Trop Dis. 2022 Jan 7;16(1):e0010019. doi: 10.1371/journal.pntd.0010019. PMID: 34995277; PMCID: PMC8797211.

Lindenbach, B.T., C.L. Murray, H.-J. Thiel, and C. Rice. 2013. Flaviviridae. In Fields Virology. D.M. Knipe and P.M. Howley, editors. Lippincott Williams & Wilkins, Philadelphia. 712–746. medRxiv 2022.11.04.22281958; doi: https://doi.org/10.1101/2022.11.04.22281958.

Minas Gerais State Department of Health. 2022. Febre Amarela. Avaialble in: https://www.saude.mg.gov.br/febreamarela.

Marengo Jose A., Ambrizzi Tercio, Alves Lincoln M., Barreto Naurinete J. C., Simões Reboita Michelle, Ramos Andrea M. (2020) Changing Trends in Rainfall Extremes in the Metropolitan Area of São Paulo: Causes and Impacts Frontiers in Climate doi.org/10.3389/fclim.2020.00003

Monath, T.P., and P.F.C. Vasconcelos. 2015. Yellow fever. J. Clin. Virol. 64:160–173. doi:10.1016/j.jcv.2014.08.030.

Moreno ES, Agostini I, Holzmann I, Bitetti MSD, Oklander LI, Kowalewski MM, Beldomenico PM, Goenaga S, Martínez M, Lestani E, Desbiez ALJ, Miler P. 2015. Yellow fever impact on brown howler Monkeys (Alouatta guariba clamitans) in Argentina: a metamodelling approach based on population viability analysis and epidemiological dynamics. Memórias do Instituto Oswaldo Cruz 110: 865–876. https://www.ncbi.nlm.nih.gov/pmc/articles/PMC4660615/

National Institute of Spatial research, 2022. Report available at: https://queimadas.dgi.inpe.br/queimadas/portal-static/estatisticas_estados/.

Nguyen, L. T., Schmidt, H. A., von Haeseler, A. & Minh, B. Q. IQ-TREE: a fast and effective stochastic algorithm for estimating maximum-likelihood phylogenies. Mol. Biol. Evol. 32, 268–274 (2015).

Obolski, U. et al. MVSE: an R-package that estimates a climate-driven mosquitoborne viral suitability index. Methods Ecol. Evol. 10, 1357–1370 (2019).

Possas C, Lourenço-de-Oliveira R, Tauil PL, Pinheiro FP, Pissinatti A, Cunha RVD, Freire M, Martins RM, Homma A. Yellow fever outbreak in Brazil: the puzzle of rapid viral spread and challenges for immunisation. Mem Inst Oswaldo Cruz. 2018 Sep 3;113(10):e180278.

Pybus OG, Suchard MA, Lemey P, Bernardin FJ, Rambaut A, Crawford FW, Gray RR, Arinaminpathy N, Stramer SL, Busch MP, Delwart EL. 2012. Unifying the spatial epidemiology and molecular evolution of emerging epidemics. Proc Natl Acad Sci U S A 109:15066–15071. https://doi.org/10.1073/pnas.1206598109.

Quick J, Grubaugh ND, Pullan ST, Claro IM, Smith AD, Gangavarapu K, Oliveira G, Robles-Sikisaka R, Rogers TF, Beutler NA, Burton DR, Lewis-Ximenez LL, de Jesus JG, Giovanetti M, Hill SC, Black A, Bedford T, Carroll MW, Nunes M, Alcantara LC, J., Sabino EC, Baylis SA, Faria NR, Loose M, Simpson JT, Pybus OG, Andersen KG, Loman NJ. 2017. Multiplex PCR method for MinION and Illumina sequencing of Zika and other virus genomes directly from clinical samples. Nat Protoc 12:1261–1276. https://doi.org/10.1038/nprot.2017.066.

Rambaut A, Drummond AJ, Xie D, Baele G, Suchard MA. 2018. Posterior summarization in Bayesian phylogenetics using Tracer 1.7. Syst Biol 67:901–904. https://doi.org/10.1093/sysbio/syy032.

Rambaut, A., Lam, T. T., Max Carvalho, L. & Pybus, O. G. Exploring the temporal structure of heterochronous sequences using TempEst (formerly Path-O-Gen). Virus Evol. 2, vew007 (2016).

Regoto, P., Dereczynski, C., Chou, S. C., & Bazzanela, A. C. (2021). Observed changes in air temperature and precipitation extremes over Brazil. International Journal of Climatology, 41(11), 5125–5142. doi:10.1002/joc.7119

Ribeiro SP, Vale MM, Diniz-Filho JAF, Fernandes GW, Reis AB, Grelle CEV. 2022. Heading back into the perfect storm: increasing risks for disease emergence in Brazil? Journal of the Brazilian Society of Tropical Medicine 55: e0640–2021

Romano, A., Z. Costa, D.G. Ramos, M.A. Andrade, and J. Vds. 2014. Yellow Fever Outbreaks in Unvaccinated Populations, Brazil. PLoS Negl Trop Dis. 8:2740. doi:10.1371/journal.pntd.0002740.

Romano Alessandro Pecego Martins; Ramos, Daniel Garkauskas; Araújo, Francisco Anilton Alves; Siqueira, Giselle Angélica Moreira de; Ribeiro, Mariana Pelissari Dias; Leal, Silvana Gomes; Elkhoury, Ana Nilce Maia Silveira. Yellow fever in Brazil: recommendations for surveillance, prevention and control. Epidemiol. serv. saúde ; 20(1): 101–106, 2011

Sacchetto L, Drumond BP, Han BA, Nogueira ML, Vasilakis N. Re-emergence of yellow fever in the neotropics - quo vadis? Emerg Top Life Sci. 2020 Dec 11;4(4):399–410. doi: 10.1042/ETLS20200187. PMID: 33258924; PMCID: PMC7733675.

Schneider C, Coudel E, Cammelli F, Sablayrolles P. 2015. Small-scale farmers’needs to end deforestation: insights for REDD+ in São Felix do Xingu (Pará, Brazil). International Forestry Review 17S1: 124–142.https://www.jstor.org/stable/26431590

Silva, N.I.O., L. Sacchetto, I.M. De Rezende, G.D.S. Trindade, A.D. Labeaud, B. De Thoisy, and B.P. Drumond. 2020. Recent sylvatic yellow fever virus transmission in Brazil: The news from an old disease. Virol. J. 17. doi:10.1186/s12985-019-1277-7.

Suchard, M. A. et al. Bayesian phylogenetic and phylodynamic data integration using BEAST 1.10. Virus Evol. 4, 1–5 (2018).

Taishi Nakase, Marta Giovanetti, Uri Obolski, José Lourenço. medRxiv 2022.11.04.22281958; doi: https://doi.org/10.1101/2022.11.04.22281958

Tisler TR, Teixeira FZ, Nóbrega RAA. Conservation opportunities and challenges in Brazil’s roadless and railroad-less areas. Sci Adv. 2022 Mar 4;8(9):eabi5548. doi: 10.1126/sciadv.abi5548. Epub 2022 Mar 4. PMID: 35245118; PMCID: PMC8896799.

Tuboi SH, Costa ZG, da Costa Vasconcelos PF, Hatch D. Clinical and epidemiological characteristics of yellow fever in Brazil: analysis of reported cases 1998-2002. Transactions of the Royal Society of Tropical Medicine and Hygiene. 2007; 101:169–175. [PubMed: 16814821]

Vasconcelos, P.F., Rodrigues, S.G., Degallier, N., Moraes, M.A., da Rosa, J.F., da Rosa, E.S. et al. (1997) An epidemic of sylvatic yellow fever in the southeast region of Maranhao State, Brazil, 1993–1994: epidemiologic and entomologic findings. Am. J. Trop. Med. Hyg. 57, 132–137 https://doi.org/10.4269/ajtmh.1997.57.132

Vasconcelos, P.F.C., J.E. Bryant, A.P.A. Travassos Da Rosa, R.B. Tesh, S.G. Rodrigues, and A.D.T. Barrett. 2004. Genetic Divergence and Dispersal of Yellow Fever Virus, Brazil. Emerg. Infect. Dis. 10:1578–1584.

Vasconcelos PFC, Sperb AF, Monteiro HAO, Torres MAN, Sousa MRS, Vasconcelos HB, et al. Isolations of yellow fever virus from Haemagogus leucocelaenus in Rio Grande do Sul State, Brazil. Trans R Soc Trop Med Hyg. 2003;97:60–2.

Vilsker, M. et al. Genome detective: an automated system for virus identification from high-throughput sequencing data. Bioinformatics 35, 871–873 (2019).

