## Supplementary_Appendix for "Genomic epidemiology sheds light on the recent spatio-temporal dynamics of Yellow Fever virus and the spatial corridor that fueled its ongoing emergence in southern Brazil"

**Supplementary Material**

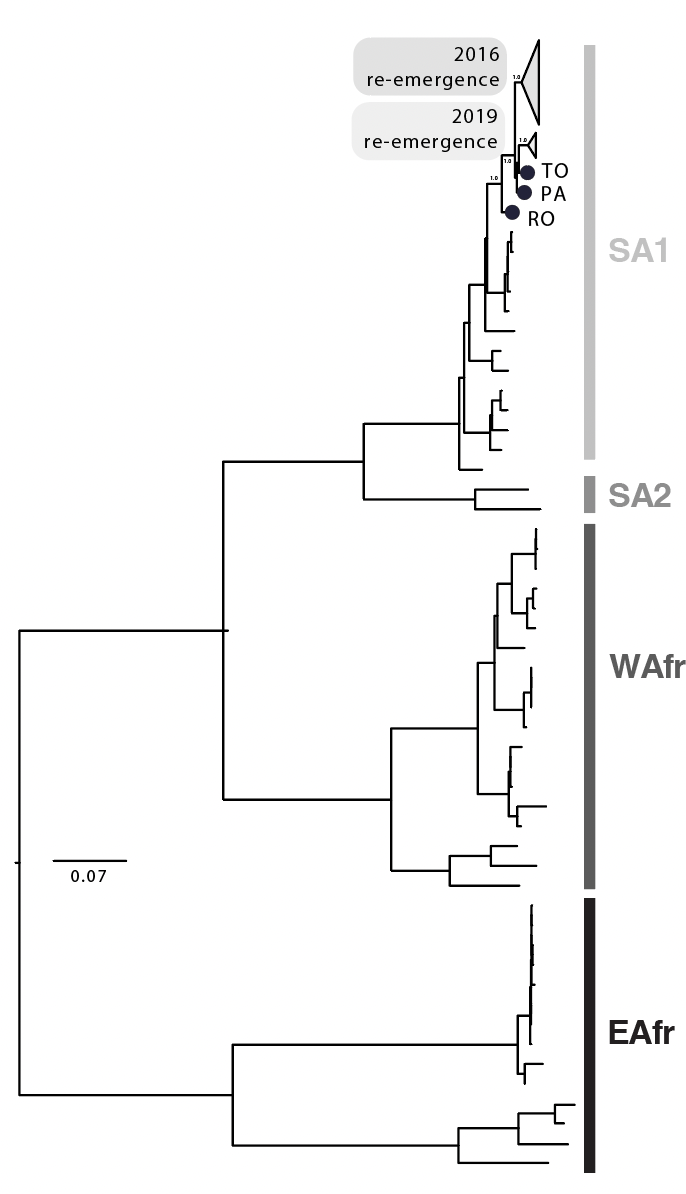

**Figure S1. Evolutionary history of the Brazilian YFV epidemics.** Maximum likelihood phylogeny of complete YFV genomes showing the outbreak clades (gray triangles) within the South American I (SA1) genotype. The scale bar is in units of substitutions per site (subst/site). Node labels indicate bootstrap support values. RO indicates a strain from Roraima isolated in 2002. PA indicates a strain from Para isolated in 2017 and TO indicates a strain from Tocantins isolated in 2017.

**Supplementary Text**

### 1. Parameterization of Index P for Yellow Fever

#### 1.2 Summary

Index P is a suitability measure that quantifies the transmission potential of adult female mosquitoes in relation to a specific host-mosquito-pathogen system [[1,2]](https://paperpile.com/c/lHYMXG/MyvD+npiV). As its formulation is based on a mathematical modeling framework for the transmission of Zika virus [[3]](https://paperpile.com/c/lHYMXG/Q6tm), the index takes into consideration the existence of a single host and a single vector. In recent years, it has mostly been used to study the transmission dynamics of mosquito-borne viruses among humans and *Aedes sp*. mosquitoes, such as dengue, Zika and chikungunya (e.g. [[4]](https://paperpile.com/c/lHYMXG/os2s) [[1,4,5]](https://paperpile.com/c/lHYMXG/os2s+1Tgm+MyvD)). However, it has also been recently used to estimate the risk of spillover of West Nile virus (WNV) [[6–8]](https://paperpile.com/c/lHYMXG/Np1W+Yb4b+DkEz), a zoonotic pathogen maintained in a transmission cycle among multiple species of mosquito and (mainly) avian species. In such a case, the modeled host is an aggregated (average) avian species taking central role in the reservoir transmission cycle (rather than the human host), and the modeled vector is the mosquito species responsible for the spillover events (rather than all the mosquito species involved in the transmission cycle in the animal reservoir). In this study we take a similar approach to that applied to WNV, modeling an ideal (average) non-human primate (NHP) host and the mosquito *Aedes aegypti* (as the main vector of YFV within the urban cycle of transmission and likely a vector for spillover events into human populations). As such, we interpret Index P as a proxy for the risk of spillover from the animal reservoir via the urban vector-species.

The index P receives as input climatic variables (humidity, temperature) and a few user-defined probability distributions related to host-virus-vector traits. We used satellite climate data from Copernicus.eu (dataset “ERA5-Land monthly averaged data from 1950 to present”, <https://cds.climate.copernicus.eu/>), and parameter probability distributions informed by the literature (Table S1). We used MVSE R-package v1.01 to estimate index P [[1]](https://paperpile.com/c/lHYMXG/MyvD).

For completeness, equation 1 presents the mathematical expression of the index P. For the mathematical expressions of each climate-dependent parameter, please refer to the full methodological description by Obolski et al. [[1]](https://paperpile.com/c/lHYMXG/MyvD). Table S1 presents a summary description of the parameters as well as the user-defined probability distributions related to YFV, NHP and *Aedes aegypti*.

Equation 1: mathematical expression of index P (U is relative air humidity and T is temperature)

$P_{(U,T)}=\frac{a^{v}(U)^{2} \Phi^{v\to h}(T) \Phi^{h\to v}(T) \gamma^{v}(T) \gamma^{h}}{\mu^{v}(U,T) (\sigma^{h}+\mu^{h}) (\gamma^{h} + \mu^{h}) (\gamma^{v}(T) + \mu^{v}(U,T))}$

**Table S1.** Proposed probability distributions related to the host-pathogen system under study for MVSE R-package input

| Parameter | Symbol | Type | Notes | References |
| --- | --- | --- | --- | --- |
| Mosquito lifespan | $1/\mu^{v}$ | $Normal(\mu,\sigma^{2})$, $\mu=12$, $\sigma=2$  (days) | *Aedes aegypti*; used in original Index P formulation. | [[1,9,10]](https://paperpile.com/c/lHYMXG/MyvD+4xvT+Bgpz) |
| Mosquito (extrinsic) incubation period | ${1/\gamma}^{v}$ | $Normal(\mu,\sigma^{2})$, $\mu=26$, $\sigma=4.6$  (days) | YFV; *Aedes aegypti* **^∗^** | See below. |
| Mosquito biting rate | $a^{v}$ | 0.25 per day | *Aedes aegypti*; used in original Index P formulation. | [[11]](https://paperpile.com/c/lHYMXG/CA2w) |
| Host lifespan | ${1/\mu}^{h}$ | $Normal(\mu,\sigma^{2})$, $\mu=17$, $\sigma=2$  (years) | (average) non-human primate **^∞^** | See below. |
| Host (intrinsic) incubation period | ${1/\gamma}^{h}$ | $Normal(\mu,\sigma^{2})$, $\mu=3$, $\sigma=0.5$  (days) | YFV; Rhesus macaques **^∗^** | See below. |
| Host (intrinsic) infectious period | ${1/\sigma}^{h}$ | $Normal(\mu,\sigma^{2})$, $\mu=4$, $\sigma=0.5$  (days) | YFV; Rhesus macaques **^∗^** | See below. |
| Probability of transmission per bite from human to mosquito | $\Phi^{h\to v}$ | MVSE R-package does not receive a distribution for this parameter. | DENV; *Aedes aegypti;* used in original Index P formulation. ∎ | See below. |
| Probability of transmission per bite from mosquito to human | $\Phi^{v\to h}$ | MVSE R-package does not receive a distribution for this parameter. | DENV; *Aedes aegypti*; used in original Index P formulation. ∎ | See below. |

^∗^ Description in sections 1.3 and 1.4 below.

**^∞^** Description in section 1.5 below.

∎ Description in section 1.6 below.

**1.3 Extrinsic incubation period (EIP)**

The probability distribution of the EIP for YFV transmitted by *Aedes aegypti* is based on the study by Johansson et al. 2010 [[12]](https://paperpile.com/c/lHYMXG/MI4Z). It is assumed that the EIP ($1/\sigma^{v}(T)$) is exponentially distributed and that it is temperature-dependent according to $\sigma^{v}(T) = 1/e^{(\beta_{0}+\beta_{1}T)}$, where $\sigma^{v}$ is the rate of becoming infectious and $T$ the temperature in Celsius. As such, $1/\sigma^{v}(T)$ is the mean EIP at a fixed temperature $T$. Johansson et al. provide uncertainty estimates for $\beta_{0}=4.6$ and $\beta_{1}=-0.07$. The mean annual temperatures in the regions of study in Brazil range approximately between 15 and 25 Celsius, such that the mean EIP would range between 17 and 35 days. For simplicity, we formalize a normal probability distribution where 95% of its density is between 17 and 35 days, such that the mean EPI has the distribution $Normal(\mu,\sigma^{2})$ with $\mu=26$ and $\sigma=4.6$ days (where $\mu$ is the mean and $\sigma$ the standard deviation).

**1.4 Intrinsic incubation and infectious periods**

Most observed cases of YFV in the region of study occurred among *Alouatta sp.,* followed by *Callithrix sp.*, *Sapajus sp.*, *Cebus sp.* and *Leontopithecus sp.*. There is a lack of high resolution data on the course of infection of YFV in such non-human primate species, with most existing data onthe intrinsic incubation period (IIP) drawn fromstudies performed on Rhesus macaques in the early 1900s. For example, Hindle (1930) notes an incubation period of 2 to 5 days in Rhesus macaques with host infectivity stopping between 1 to 3 days after the onset of fever [[12,13]](https://paperpile.com/c/lHYMXG/MI4Z+adI9). Beeuwkes (1936) on the other hand reported an incubation period lasting 6 days after inoculation of a single Rhesus macaque with blood from a human infected with YFV [[14]](https://paperpile.com/c/lHYMXG/1Z7Y). Hudson and Phillip (1929) also infected five Rhesus macaques with YFV, of which three were infected by a mosquito bite [[15]](https://paperpile.com/c/lHYMXG/UQLb): one macaque was infectious for 4 days with an incubation period and latent period of 2 days and 1 day respectively; a second macaque was infectious for 5 days with an incubation period and latent period of 2 days and 1 day respectively; a third monkey was infectious for 4 days with an incubation period and latent period of 2 days and 1 day respectively. From such limited data, we assume with simplicity that the IIP follows the distribution $Normal(\mu,\sigma^{2})$ with $\mu=3$ and $\sigma=0.5$ days (where $\mu$ is the mean and $\sigma$ the standard deviation), and the non-human primate infectious period follows the distribution $Normal(\mu,\sigma^{2})$ with $\mu=4$ and $\sigma=0.5$ days (where $\mu$ is the mean and $\sigma$ the standard deviation).

**1.5 Host lifespan**

We searched the Human Ageing Genomic Resources (HAGR, [[16]](https://paperpile.com/c/lHYMXG/otT9)) website that contains a database on animal species longevity. The database describes that *Alouatta sp.* can reach ages >20 years in captivity, although a few more specific examples such as that for *Alouatta palliata* show that individuals in the wild typically live to 16 years of age. For *Callithrix sp.* the database reports longevity of 15 to 25 years for captive individuals, which is likely shorter for wild individuals. Given that most YFV infections observed were among *Alouatta and Callithrix sp.*, we defined host lifespan as $Normal(\mu,\sigma^{2})$ with $\mu=17$ and $\sigma=2$ years (where $\mu$ is the mean and $\sigma$ the standard deviation).

**1.6 Probability of transmission per infectious bite**

Due to lack of existing data for the modelled system related to YFV, we assume the probabilities $\Phi^{v\to h}(T)$ (mosquito-to-human) and $\Phi^{h\to v}(T$) (human-to-mosquito) to behave similarly to DENV in relation to temperature for *Aedes aegypti*. For the mathematical expressions underlying these parameters please refer to the full methodological description by Obolski et al. [[1]](https://paperpile.com/c/lHYMXG/MyvD).

### 2. Bayesian spatial regression

#### 2.1 Summary of the modelling framework

Our spatial model for YFV case counts is based on the Besag York Mollié (BYM2) model [[17,18]](https://paperpile.com/c/lHYMXG/SP43+6NPR), which assumes cases counts $n_{i}$ in municipality $i=1,\ldots,N$ to be Poisson distributed with mean:

$\mu_{i}= N_{i}exp(\alpha+\sum_{j} \omega_{j}X_{ij}+\psi_{i})$ ,

where $N_{i}$ is population size, $\alpha$ is the intercept term, $X$ is a matrix of spatial covariates, the $\omega$'s represent fixed effects and the $\psi$'s are random effects accounting for unobserved variation. Here we follow the parametrization as in [[19]](https://paperpile.com/c/lHYMXG/44Ik), where the random effects are parameterized as:

$\psi=\sigma(\sqrt{1-\rho} {\cdot\theta}+\sqrt{\rho}\cdot\phi' )$ ,

Where $\theta$, $\phi'$ represent spatially unstructured and structured components, respectively. The parameter $\rho\in[0,1]$ quantifies the relative contribution of spatial variation with respect to unstructured variation. $\theta$, $\phi'$ are assumed to have approximately unit variance, so that the overall variance of random effects is $\sigma^{2}$. For this to be achieved, $\phi'$ is defined as $\phi/\sqrt{s}$, where $s=0.51$ is a scaling factor depending solely on the geometry of the study area [[19]](https://paperpile.com/c/lHYMXG/44Ik). $\phi$ is an intrinsic conditional autoregressive (CAR) component accounting for spatial autocorrelation between neighboring locations, and is defined by the conditional multivariate normal distribution:

$\phi_{i}|\phi_{j},j\neq i \sim Normal(d_{i}^{-1}\sum_{j\sim i} \phi_{j},d_{i}^{-1}) ,$

where the notation $j\sim i$ includes the $d_{i}$ neighbors of location $i$ only. We also add a soft constraint on $\phi$ so that $\sum_{i}^{N} \phi_{i}\approx0$, lifting non identifiability in $\phi$. A detailed account of priors can be found in Table S2.

**Table S2.** Summary of parameters and their prior distributions for the spatial regression analysis.

| Parameter | Symbol | Prior | Notes |
| --- | --- | --- | --- |
| Intercept | $\alpha$ | $Normal(0,1)$ | ---- |
| Fixed effects weights | $\omega$ | $Normal(0,1)$ | ---- |
| Random effect scale | $\sigma$ | $Exponential(1)$ | ---- |
| Relative weight of spatial effects | $\rho$ | $ln\frac{\rho}{1-\rho}\sim Normal(0,1)$ | ---- |
| Non-spatial effects | $\theta$ | $Normal(0,1)$ | ---- |
| Spatial effects | $\phi$ | $ICAR$ prior, with  $\sum_{i}^{N} \phi_{i}\sim Normal(0,N/1000)$ | The soft prior on $\sum_{i}^{N} \phi_{i}$  is for identifiability |

**2.2 Details on covariate data**

We obtained estimates of vaccination coverage for 2016 from [[20]](https://paperpile.com/c/lHYMXG/yEE8). The authors provided three estimates for YFV vaccination coverage, based on different assumptions about vaccine allocation. Here, we used their most conservative estimate, obtained under the assumption of unbiased and untargeted vaccine allocation. In the spatial regression framework, we used cumulative counts per municipality up to 2019, scaled by the largest value in the sample. We calculated Mean Index P monthly for each municipality using the procedure outlined in Section 1. Index P values from 2016 to 2020 were then averaged together to yield a single value per municipality. Forest cover, defined as the fraction of ground covered by green vegetation, was obtained from [[21]](https://paperpile.com/c/lHYMXG/qEQ5). We obtained three values corresponding to March, July and November 2016; these were then averaged into a single value per municipality. Other covariates, i.e. mean altitude, population density (relative to 2015), extent of pastures and urban areas and extent of Atlantic Forest and Cerrado biomes, were obtained from [[22]](https://paperpile.com/c/lHYMXG/HQ5o). In the fit, these variables were scaled by the corresponding maximum value in the sample. Summary of variables is in **Table S3**.

**Table S3.** Summary of covariate data

| Variable | Definition | Form/Type | Source |
| --- | --- | --- | --- |
| Atlantic | Biome classification of Atlantic forest type. | Mean of 2006 per municipality. $ | [[22]](https://paperpile.com/c/lHYMXG/HQ5o) |
| NHP/YFV count | Number of NHP YFV infections. | Cumulative sum per municipality, up to 2019. | Dataset reported in this study. |
| Forest cover | Fraction of ground covered by green vegetation. | Mean of 3 monthly timepoints in 2016 per municipality. | [[21]](https://paperpile.com/c/lHYMXG/qEQ5) |
| Altitude | Land altitude. | Mean of 2000 per municipality. $ | [[22]](https://paperpile.com/c/lHYMXG/HQ5o) |
| Pasture | Land use classification: predominantly planted, linked to agricultural activity. | Mean of 2018 per municipality. $ | [[22]](https://paperpile.com/c/lHYMXG/HQ5o) |
| Cerrado | Biome classification of Cerrado vegetation type. | Mean of 2006 per municipality. $ | [[22]](https://paperpile.com/c/lHYMXG/HQ5o) |
| Urbanization | Land use classification: predominance of  non-vegetated surfaces, including roads, roads and  buildings. | Mean of 2018 per municipality. $ | [[22]](https://paperpile.com/c/lHYMXG/HQ5o) |
| Vaccination coverage | Estimates of vaccination coverage. | Most conservative mean estimation for 2016 per municipality. | [[20]](https://paperpile.com/c/lHYMXG/yEE8) |
| Population density | Human population density. | Mean of 2015 per municipality. ₳ $ | [[22]](https://paperpile.com/c/lHYMXG/HQ5o) |
| Index P | Risk of spillover from the animal reservoir via the urban vector-species. | Mean of 2016-2020 per municipality. | Estimated in this study. |

#### $ as stated in the original data set [[22]](https://paperpile.com/c/lHYMXG/HQ5o): “Mean area of the features/geometries or of the values for each class or variable in the thematic layers contained in each municipality”.

₳ as stated in the original data set [[22]](https://paperpile.com/c/lHYMXG/HQ5o): “Demographic density data (measure expressed by the relationship between the population and the area of the territory) interpolated”.

#### 2.3 Details on model implementation and fitting

For some covariates, missing values in a few municipalities were imputed using average values from immediate neighbors. We considered two municipalities to be neighbors if they shared a border using geopandas module v0.11.1 [[23]](https://paperpile.com/c/lHYMXG/2oZQ). We excluded a single municipality, Ilhabela, for which it was problematic to assign neighbors. The model was implemented in Stan [[23,24]](https://paperpile.com/c/lHYMXG/2oZQ+KT1b) and run in R v4.1.3 through the rstan interface v2.21.7 [[23–25]](https://paperpile.com/c/lHYMXG/2oZQ+KT1b+c6X8). We ran 4 independent chains and for each we collected 4000 posterior samples after a warmup of 2000 steps. MCMC diagnostics indicated no divergent transitions during sampling. Also, the R-hat convergence diagnostic was less than 1.05, suggesting convergence. Model output is presented in **Figure S2**, estimations in **Table S4**.

**Table S4.** Regression weights from spatial modelling

| Variable | Median | Effect | 95% range | Significant * |
| --- | --- | --- | --- | --- |
| Atlantic | 0.79 | Positive | -0.28, 1.89 | No |
| NHP/YFV count | 0.74 | Positive | -0.49, 1.99 | No |
| Forest cover | 0.26 | Positive | -1.25, 1.72 | No |
| Altitude | 0.07 | Positive | -1.14. 1.27 | No |
| Pasture | -0.03 | Negative | -1.39, 1.32 | No |
| Cerrado | -0.48 | Negative | -2.32, 1.29 | No |
| Urbanization | -0.52 | Negative | -2.42, 1.31 | No |
| Vaccination coverage | -1.09 | Negative | -2.28, 0.03 | No |
| Population density | -3.23 | Negative | -4.78, -1.65 | Yes |
| Index P | -5.63 | Negative | -7.37, -3.86 | Yes |

* If the 95% range of the posterior included zero or not.

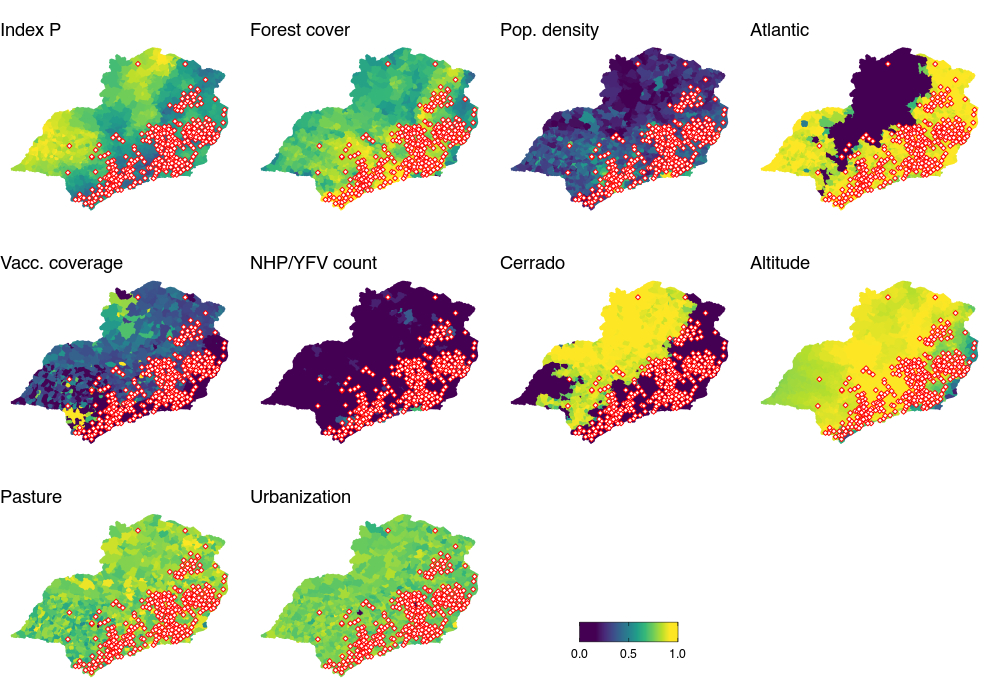

**Figure S2. Spatial distribution of ten covariates and NHP infections.** Maps present the covariates per municipality (see **Table S3**). All covariates were normalised by their maximum for visualisation, with covariates population density, urbanization, Atlantic, Cerrado, and NHP cases first transformed with log10. The red points mark the locations of the reported human YFV infections.

| 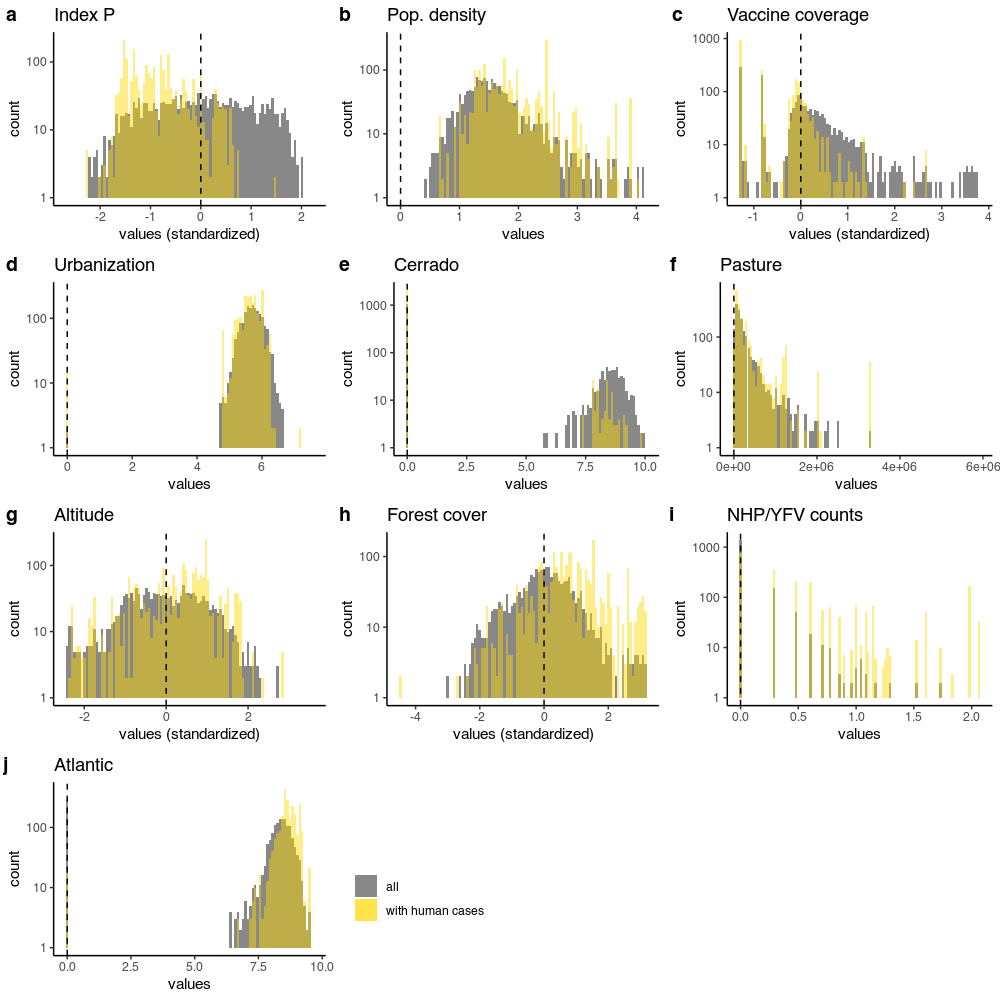 | **Figure S3. Relationships of human cases versus covariates used for spatial modelling.**  (a) Distribution of all observed values of index P (grey) across the spatial range of the study (municipalities), and observed values of index P for municipalities presenting human YFV cases (yellow). (b-j) Same as in panel a but for the other covariates. (a-j) Values are presented for the Southeast and South macroregions of Brazil. For visualization, some variables were standardized while others were transformed with log10 (see x-axis titles). |
| --- | --- |

| 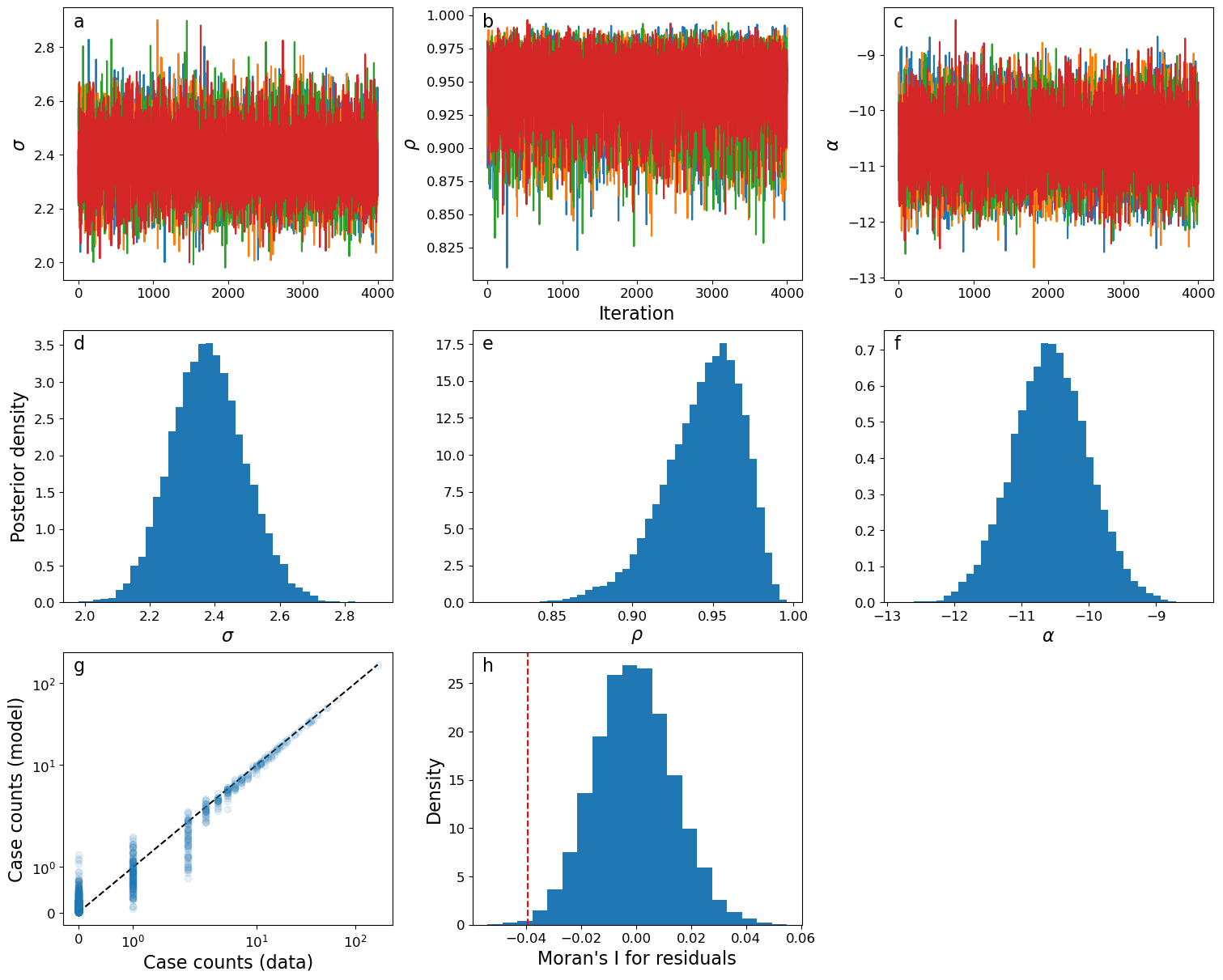 | **Figure S4.** **Trace plots for spatial regression modelling.**  (a-c) Trace plots for parameters $\sigma$, $\rho$ and $\alpha$. Each line corresponds to a different chain (d-f) Posterior marginal distributions. (g) Fitted versus observed case counts. (h) Spatial autocorrelation of model residuals. We quantify spatial autocorrelation among mean posterior residuals by calculating Moran's I statistic using the same area adjacency matrix used in the main analysis [[26]](https://paperpile.com/c/lHYMXG/GqSZ). The value of Moran's I obtained directly from residuals (line) is compared with a null distribution obtained by permuting residuals 10000 times. Panel h shows absence of positive autocorrelation albeit some degree of negative spatial autocorrelation is present. |
| --- | --- |

| 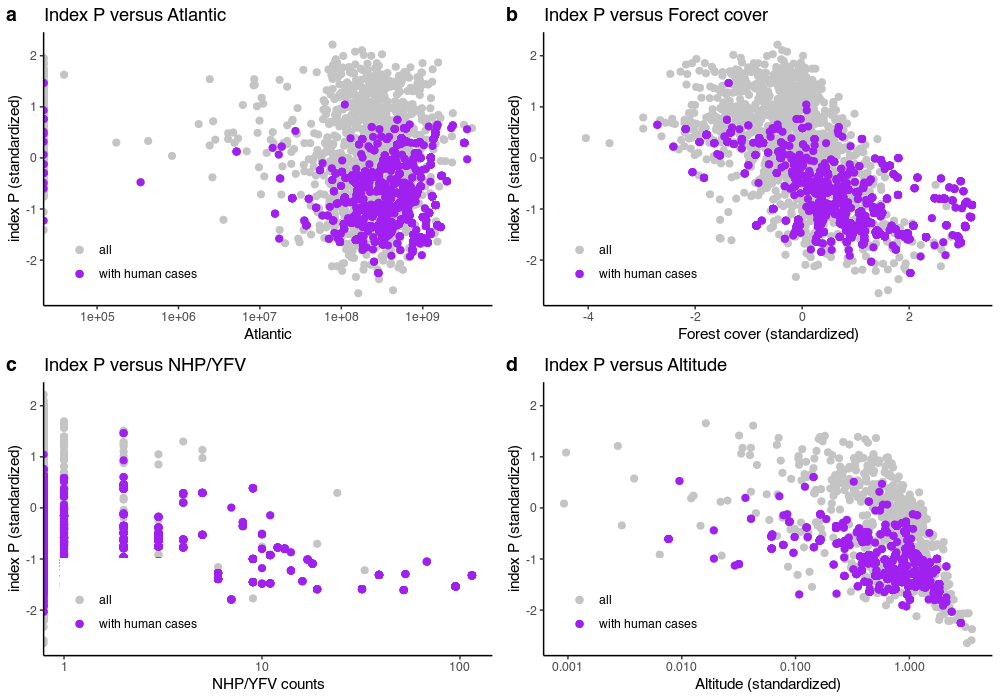 | **Figure S5. Scatterplot of local Index P versus spatial regression covariates with median positive effect.**  Panels show the values of index P and other variables - values for all municipalities (grey) and values for municipalities with human cases (purple). Each point is a municipality within the Southeast and South macroregions of Brazil. |
| --- | --- |

| 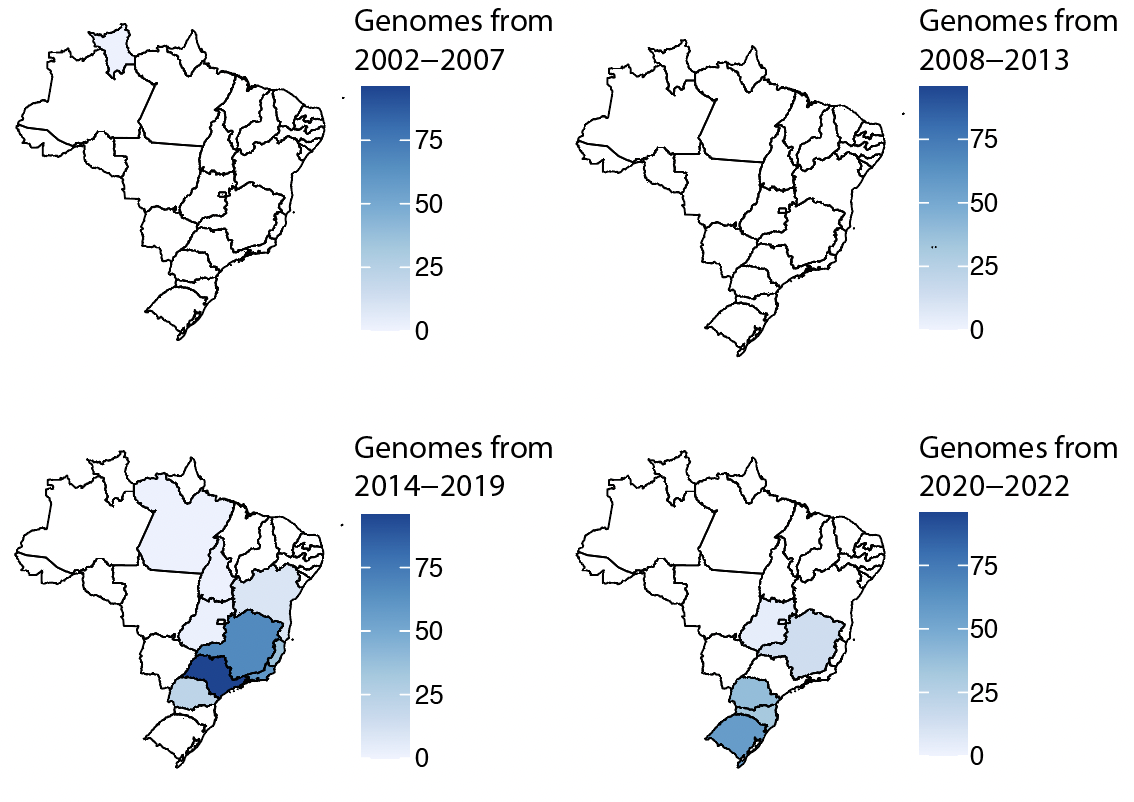 | **Figure S6.** Spatial distribution of YFV complete genomes sequences available in Public Database (NCBI) since 2002. |
| --- | --- |

#### 3. Lineage specific genetic signatures

**Table S5.** Amino acid substitutions in the YFV coding region within the clades Ia-IIIc. Amino acid (aa) abbreviations: S: Serine, H: Histidine; Q: Glutamine; D: Aspartate; G: Glycine; T: Threonine; P: Proline; V: Valine; I: Isoleucine; R: Arginine; N: Asparagine.

| **Genomic position** | **Reference** | **Mutation** | **Type of substitution** | **Amino acid changes** | **Clade** | **Genomic region** |
| --- | --- | --- | --- | --- | --- | --- |
| 2827 | T | C | Synonymous | S-S | Ia | NS1 |
| 5362 | T | C | Synonymous | H-H | Ia | NS3 |
| 6229 | G | A | Synonymous | Q-Q | Ia | NS3 |
| 8053 | A | G | Nonsynonymous | S-D | Ia | RdRp |
| 9570 | T | C | Synonymous | G-G | Ia | RdRp |
| 9806 | C | T | Nonsynonymous | T-I | Ia | RdRp |
| 8047 | A | G | Synonymous | P-P | IIb | RdRp |
| 8048 | G | A | Nonsynonymous | V-I | IIb | RdRp |
| 6184 | C | A | Synonymous | R-R | IIIc | NS3 |
| 8048 | G | A | Nonsynonymous | S-N | IIIc | RdRp |

**Supplementary References**

1.	Obolski U, Perez PN, Villabona-Arenas CJ, Thézé J, Faria NR, Lourenço J. MVSE: An R-package that estimates a climate-driven mosquito-borne viral suitability index. Methods Ecol Evol. 2019;10. doi:[10.1111/2041-210X.13205](http://dx.doi.org/10.1111/2041-210X.13205)

2.	Nakase T, Giovanetti M, Obolski U, Lourenço J. Global transmission suitability maps for dengue virus transmitted by Aedes aegypti from 1981 to 2019. medRxiv. 2022. p. 2022.11.04.22281958. doi:[10.1101/2022.11.04.22281958](http://dx.doi.org/10.1101/2022.11.04.22281958)

3.	[Lourenço J, de Lima MM, Faria NR, Walker A, Kraemer MUG, Villabona-Arenas CJ, et al. Epidemiological and ecological determinants of Zika virus transmission in an urban setting. 2017 [cited 7 Nov 2022]. doi:](http://paperpile.com/b/lHYMXG/Q6tm)[10.7554/eLife.29820](http://dx.doi.org/10.7554/eLife.29820)

4.	Adelino TÉR, Giovanetti M, Fonseca V, Xavier J, de Abreu ÁS, do Nascimento VA, et al. Field and classroom initiatives for portable sequence-based monitoring of dengue virus in Brazil. Nat Commun. 2021;12: 1–12.

5.	Petrone ME, Earnest R, Lourenço J, Kraemer MUG, Paulino-Ramirez R, Grubaugh ND, et al. Asynchronicity of endemic and emerging mosquito-borne disease outbreaks in the Dominican Republic. Nat Commun. 2021;12: 1–12.

6.	Lourenço J, Pinotti F, Nakase T, Giovanetti M, Obolski U. Letter to the editor: Atypical weather is associated with the 2022 early start of West Nile virus transmission in Italy. Euro Surveill. 2022;27. doi:[10.2807/1560-7917.ES.2022.27.34.2200662](http://dx.doi.org/10.2807/1560-7917.ES.2022.27.34.2200662)

7.	Lourenço J, Barros SC, Zé-Zé L, Damineli DSC, Giovanetti M, Osório HC, et al. West Nile virus transmission potential in Portugal. Communications Biology. 2022;5: 1–12.

8.	Lourenço J, Thompson RN, Thézé J, Obolski U. Characterising West Nile virus epidemiology in Israel using a transmission suitability index. Eurosurveillance. 2020;25: 1900629.

9.	Hugo LE, Jeffery JAL, Trewin BJ, Wockner LF, Nguyen TY, Nguyen HL, et al. Adult survivorship of the dengue mosquito Aedes aegypti varies seasonally in central Vietnam. PLoS Negl Trop Dis. 2014;8: e2669.

10.	Trpis M, Häusermann W, Craig GB Jr. Estimates of population size, dispersal, and longevity of domestic Aedes aegypti aegypti (Diptera: Culicidae) by mark-release-recapture in the village of Shauri Moyo in eastern Kenya. J Med Entomol. 1995;32: 27–33.

11.	Trpis M, Hausermann W. Dispersal and other Population Parameters of Aedes aegypti in an African Village and their Possible Significance in Epidemiology of Vector-Borne Diseases. The American Journal of Tropical Medicine and Hygiene. 1986. pp. 1263–1279. doi:[10.4269/ajtmh.1986.35.1263](http://dx.doi.org/10.4269/ajtmh.1986.35.1263)

12.	Johansson MA, Arana-Vizcarrondo N, Erin Staples J. Incubation Periods of Yellow Fever Virus. Am J Trop Med Hyg. 2010;83: 183–188.

13.	Hindle E. THE TRANSMISSION OF YELLOW FEVER. Lancet. 1930;216: 835–842.

14.	Beeuwkes H. Clinical manifestations of yellow fever in the West African native as observed during four extensive epidemics of the disease in the Gold Coast and Nigeria∗. Trans R Soc Trop Med Hyg. 1936;30: 61–86.

15.	Hudson NP, Philip CB. INFECTIVITY OF BLOOD DURING THE COURSE OF EXPERIMENTAL YELLOW FEVER. J Exp Med. 1929;50: 583–599.

16.	Tacutu R, Thornton D, Johnson E, Budovsky A, Barardo D, Craig T, et al. Human Ageing Genomic Resources: new and updated databases. Nucleic Acids Res. 2018;46: D1083–D1090.

17.	Website. Available: <https://mc-stan.org/users/documentation/case-studies/icar_stan.html>

18.	Besag J, York J, Mollié A. Bayesian image restoration, with two applications in spatial statistics. Annals of the Institute of Statistical Mathematics. 1991. pp. 1–20. doi:[10.1007/bf00116466](http://dx.doi.org/10.1007/bf00116466)

19.	Riebler A, Sørbye SH, Simpson D, Rue H. An intuitive Bayesian spatial model for disease mapping that accounts for scaling. Stat Methods Med Res. 2016;25: 1145–1165.

20.	Shearer FM, Moyes CL, Pigott DM, Brady OJ, Marinho F, Deshpande A, et al. Global yellow fever vaccination coverage from 1970 to 2016: an adjusted retrospective analysis. Lancet Infect Dis. 2017;17: 1209–1217.

21.	Buchhorn M, Smets B, Bertels L, De Roo B, Lesiv M, Tsendbazar N-E, et al. Copernicus Global Land Service: Land Cover 100m: version 3 Globe 2015-2019: Product User Manual. Zenodo; 2020. doi:[10.5281/ZENODO.3938963](http://dx.doi.org/10.5281/ZENODO.3938963)

22.	Abdalla L, Augusto DA, Chame M, Dufek AS, Oliveira L, Krempser E. Statistically enriched geospatial datasets of Brazilian municipalities for data-driven modeling. Sci Data. 2022;9: 489.

23.	[Jordahl K, Van den Bossche J, Fleischmann M, Wasserman J, McBride J, Gerard J, et al. geopandas/geopandas: v0.8.1. 2020 [cited 7 Nov 2022]. doi:](http://paperpile.com/b/lHYMXG/2oZQ)[10.5281/zenodo.3946761](http://dx.doi.org/10.5281/zenodo.3946761)

24.	[Stan. In: stan-dev.github.io [Internet]. [cited 7 Nov 2022]. Available:](http://paperpile.com/b/lHYMXG/KT1b) <https://mc-stan.org>

25.	[RStan: the R interface to Stan. Website. In: Stan Development Team (2022). R package version 2.21.7 [Internet]. Available:](http://paperpile.com/b/lHYMXG/c6X8) <https://mc-stan.org/>

26.	Moran PAP. Notes on continuous stochastic phenomena. Biometrika. 1950;37: 17–23.
